# Beyond greenness: Greenspace morphology associates with disability prevalence among children, working-age adults, and older adults—a nationwide study

**DOI:** 10.64898/2026.07.08.26357548

**Authors:** Simin Gholami, Jiahe Bian, Keith Christensen, Louis Tassinary, Huaqing Wang

## Abstract

Greenspace has been associated with a wide range of health outcomes and conditions related to functional limitation and disability. Yet less is known about how the spatial morphology of greenspace relates to disability prevalence across different stages of the life course. This study examines associations between greenspace morphology and disability prevalence among children, working-age adults, and older adults in urban census tracts across the contiguous United States. Using national land-cover data, we quantified morphological metrics at the census-tract level, including greenspace percentage, density, mean size, connectedness, shape complexity, inter-greenspace distance, and diversity. These indicators were linked with age-specific disability prevalence obtained from the American Community Survey. Spatial lag regression models were used to account for spatial dependence while adjusting for socio-demographic and contextual characteristics. Across age groups, higher greenspace percentage was consistently associated with lower disability prevalence (children: β = −0.081, 95% CI: −0.096 to −0.066; adults: β = − 0.804, −0.858 to −0.750; older adults: β = −1.132, −1.250 to −1.013). Among children, patch density (β = −0.045, −0.061 to −0.029), mean patch area (β = −0.029, −0.040 to −0.018), connectedness (β = −0.051, −\0.069 to −0.032), diversity (β = −0.036, −0.051 to −0.020), and inter-greenspace distance (β = 0.056, 0.039 to 0.073) were all associated with disability prevalence, whereas shape complexity was not (β = 0.004, − 0.010 to 0.018). Among working-age adults, associations were observed for mean area (β = −0.023, −0.090 to −0.002), connectedness (β = −0.127, −0.243 to −0.011), shape complexity (β = −0.123, −0.174 to −0.072), diversity (β = −0.146, −0.201 to −0.091), and inter-greenspace distance (β = 0.151, 0.059 to 0.242), whereas patch density was not significantly associated with disability prevalence (β = −0.013, −0.048 to 0.022). In older adults, all examined greenspace morphology metrics showed significant associations with disability prevalence, including patch density (β = −0.445, −0.842 to −0.049), diversity (β = −0.126, −0.188 to −0.065), and inter-greenspace distance (β = 0.455, 0.409 to 0.501). Overall, the findings suggest that higher greenspace percentage, larger patch size, greater connectedness, greater diversity, and more spatially clustered greenspace distributions are associated with lower disability prevalence across the life course, although the strength and consistency of these associations varied across age groups. The study provides national-scale evidence for incorporating greenspace morphology into urban planning and public health strategies to support more inclusive and health-supportive urban environments.

## 1. Introduction

Disability represents a major and growing public health and planning challenge worldwide. Globally, an estimated 1.3 billion people experience significant functional limitations (WHO, 2024), and in the United States, more than one in four adults report at least one disability (CDC, 2023). Disability prevalence varies substantially across the life course: approximately 4.3% of U.S. children, 11.1% of working-age adults, and 39.9% of adults aged 65 years and older report functional difficulties (CDC, 2025; U.S. Census Bureau, 2021b). As populations age and chronic conditions accumulate, disability is projected to increase, amplifying demands on health systems, communities, and the built environment. Understanding environmental determinants of disability is therefore critical for urban planners and public health professionals seeking to promote equitable and age-supportive communities.

A growing body of research suggests that greenspace may shape disability risk through multiple environmental and behavioral pathways. Conceptual syntheses highlight mechanisms linking greenspace exposure to improved health, including enhanced physical activity, psychological restoration, social cohesion, and mitigation of air pollution and urban heat (Arzberger et al., 2026; Markevych et al., 2017; O’Regan et al., 2022). Through these pathways, greenspace may reduce the accumulation of chronic disease burden, frailty, cognitive decline, and functional limitations that contribute to disability. These pathways are consistent with the Disablement Process Model (Verbrugge & Jette, 1994a), which conceptualizes disability as a dynamic process shaped by interactions between individual conditions and environmental contexts. Empirical findings, however, remain mixed. Several studies report protective associations between greater greenspace exposure and lower disability prevalence or slower functional decline (Cao et al., 2023; Feng et al., 2023; Fortune et al., 2020; Peng et al., 2020), whereas others observe null or even positive associations (Vogt et al., 2015; Wong et al., 2023), indicating that contextual conditions, spatial scale, and measurement strategies may shape observed relationships.

Importantly, most studies operate greenspace primarily as quantity or vegetation density, with limited attention to spatial morphology. Emerging evidence indicates that morphological characteristics, such as connectedness, diversity, and spatial proximity, may exert distinct influences beyond total greenspace area, including via behavioral and psychological pathways (H. Wang, Gholami, et al., 2024; H. Wang & Tassinary, 2024).

Conceptually, the relationship between neighborhood environments and disability can be situated within the Disablement Process Framework, which describes disability as the downstream outcome of a dynamic progression from pathology to impairments, functional limitations, and ultimately disability (Verbrugge & Jette, 1994). Within this framework, environmental conditions are understood as contextual factors that can either accelerate or buffer the progression of functional decline. Neighborhood greenspace may influence multiple stages of this process by shaping exposure to environmental stressors, supporting physical and psychological functioning, and facilitating everyday activities that help maintain functional capacity(Dong et al., 2025; Krischke et al., 2025; Liu et al., 2023). From this perspective, disability is not solely an individual-level condition but reflects the cumulative interaction between health status and the surrounding environment over time. This framework provides a useful theoretical foundation for examining how greenspace morphology, as a modifiable feature of the built environment, may contribute to population-level patterns of disability.

Additionally, disability determinants and potentially relevant environmental pathways differ across life stages. In childhood, disability is often linked to developmental conditions and early-life environmental exposures (Halfon & Hochstein, 2002; WHO, 2023). Among working-age adults, disability is more commonly driven by injuries, chronic disease burden, and mental health conditions (CDC, 2025; Verbrugge & Jette, 1994b), whereas in older adulthood it is largely shaped by cumulative physiological decline, multimorbidity, and mobility limitations (Clarke et al., 2008). A life-course perspective, therefore, suggests that neighborhood environments may differentially influence functional health at distinct stages of life (Ben-Shlomo & Kuh, 2002). Greenspace may support children’s developmental health, mitigate stress and cardiometabolic risks among adults (Chen et al., 2025; Jiang et al., 2019; H.

Wang & Li, 2023), and promote mobility and social engagement among older populations (Markevych et al., 2017; Twohig-Bennett & Jones, 2018). Older adults, in particular, may be more sensitive to neighborhood environmental conditions due to reduced mobility and heightened vulnerability to environmental stressors (Clarke et al., 2008; Han & Wang, 2026; J. Wang et al., 2026). Yet age-stratified evidence remains limited.

Against this backdrop, national-scale evidence is needed to clarify whether and how greenspace morphology relates to disability prevalence across diverse urban contexts, and whether these associations vary by age group. To address this gap, this study investigates the association between greenspace morphology and disability prevalence at the census-tract level across urban areas of the contiguous United States. We examine whether greenspace morphological attributes, including percentage, density, mean area, connectedness, shape complexity, spatial distantness, and diversity, are associated with disability prevalence beyond overall greenspace percentage, and whether these relationships differ among children (5–17 years), working-age adults (18–64 years), and older adults (65+ years).

## 2. Methods

### 2.1 Study area and prevalence of disability among different age groups

This cross-sectional ecological study examined census tracts across the contiguous United States, including the 48 states and the District of Columbia, as the spatial unit of analysis. Census tracts are relatively permanent statistical subdivisions of counties designed to represent neighborhood-scale populations that are relatively homogeneous in demographic and socioeconomic characteristics, typically containing between 1,200 and 8,000 residents with a mean population of approximately 4,000 (U.S. Census Bureau, 2022). They are widely used in population health and urban planning research because they provide a consistent and policy-relevant geographic scale for examining spatial variation in environmental exposures and health outcomes. To focus on developed and urbanized environments where greenspace morphology is most relevant to neighborhood context, we excluded census tracts with fewer than 425 housing units per square mile, ensuring that the analytical sample reflected predominantly urban settings. This cutoff follows the U.S. Census Bureau’s urban-core threshold and helped ensure that the analytical sample represented predominantly urban settings (U.S. Census Bureau, 2023).

Age-specific disability prevalence for the study year 2023 was obtained from the 2023 American Community Survey (ACS) 5-year estimates at the census tract level and accessed using the *tidycensus* package in R (Walker, 2024). The ACS defines disability based on reported difficulty across six functional domains: hearing, vision, cognitive functioning, ambulatory ability, self-care, and independent living, and classify individuals who report limitations in one or more domains as having a disability.

Because certain disability domains are not consistently assessed for children under age five in the ACS disability sequence (U.S. Census Bureau, 2021a), analyses for the child population were restricted to ages 5–17 years to ensure measurement comparability and reliability. For each census tract, disability prevalence was operationalized as the percentage of individuals within a given age group reporting at least one disability. Analyses were conducted separately for children (5–17 years), working-age adults (18–64 years), and older adults (65 years and older) to account for distinct etiologies and life-course trajectories of functional limitation. Census tracts with missing disability estimates were excluded from the analysis.

### 2.2 Greenspace data and image classification

Land cover data were obtained from the 2023 Annual National Land Cover Database (NLCD) Collection 1.0 for the conterminous United States, produced by the U.S. Geological Survey (USGS). The NLCD provides 30-m spatial-resolution land-cover classifications derived from Landsat imagery and processed using a standardized national classification framework (USGS, 2023). The 2023 dataset corresponds to the study year and represents the most recent nationally consistent land cover product available at the time of analysis. Land cover classes were reclassified into greenspace and non-greenspace categories based on the official NLCD legend (MRLC, 2023). Greenspace was defined as vegetated land cover types that, within developed landscapes, plausibly function as ecologically and socially relevant green environments. These included Developed Open Space; Deciduous Forest, Evergreen Forest, and Mixed Forest; Shrub/Scrub; Grassland/Herbaceous; Pasture/Hay; Cultivated Crops; Woody Wetlands; and Emergent Herbaceous Wetlands. All remaining land cover classes, including Developed Low, Medium, and High Intensity; Barren Land; and Open Water, were classified as non-greenspace. Raster processing, reclassification, and spatial extraction were conducted in ArcGIS Pro (Esri, Redlands, CA), and greenspace raster maps were spatially aligned with census tract boundaries to calculate landscape morphology metrics.

### 2.3 Quantification of greenspace morphology

To more accurately represent neighborhood-level exposure to surrounding vegetation and reduce potential boundary bias, we generated a 0.5-mile (approximately 800 m) Euclidean buffer around the boundary of each census tract. This distance approximates a 10-minute walking radius and is widely adopted in environmental health and urban planning research to reflect realistic spatial exposure to neighborhood environments (H. Wang, Huang, et al., 2024; H. Wang & Tassinary, 2019). Although network-based buffers were considered, Euclidean buffers were selected to capture the broader environmental context of vegetation surrounding residential areas rather than route-constrained accessibility. Environmental influences such as visual exposure to greenery, microclimatic regulation, air pollution mitigation, and noise attenuation may operate independently of direct park visitation and therefore extend beyond transportation networks (Markevych et al., 2017; Xu & Wang, 2025). The 0.5-mile threshold is consistent with prior tract-level studies examining the relationship between greenspace and health outcomes in the United States (Lasky et al., 2023; Wang & Tassinary, 2019) and aligns with commonly referenced planning standards.

Within each buffered tract, greenspace morphology was quantified using established landscape ecology metrics that characterize both composition and spatial configuration. Metric selection was guided by the objective of examining structural attributes of greenspace beyond total quantity, as landscape morphology may influence ecological functioning and health-relevant exposures independent of overall coverage.

Seven indicators were calculated to capture complementary dimensions of landscape structure. Greenspace percentage (PLAND) represented proportional vegetated coverage, while patch density (PD) captured landscape density by representing the number of discrete greenspace patches per unit area. Area-weighted mean patch area (AREA_MN) reflected average patch size, and patch cohesion (COHESION) assessed structural connectedness among patches. Mean Euclidean nearest neighbor distance (ENN_MN) quantified spatial proximity, area-weighted mean shape index (SHAPE_AM) characterized boundary complexity, and Shannon’s Diversity Index (SHDI) measured overall greenspace compositional diversity by capturing the variety and relative distribution of greenspace classes within each buffered tract. In this study, SHDI was calculated using all vegetated land-cover categories defined as greenspace, including Developed Open Space; forest types (Deciduous, Evergreen, and Mixed Forest); Shrub/Scrub and Dwarf Scrub; Grassland/Herbaceous; Pasture/Hay; Cultivated Crops; Woody Wetlands; and Emergent Herbaceous Wetlands. Together, these metrics describe how greenspace is distributed, configured, and structurally organized within the neighborhood landscape. All metrics were computed using standard landscape ecology formulations (Table 1), with detailed mathematical definitions referenced from Fragstats user manual (McGarigal, 2014; McGarigal et al., 2012), and calculated via *landscapemetrics* package in R (Hesselbarth et al., 2019; R Core Team, 2021).

**Table 1.**
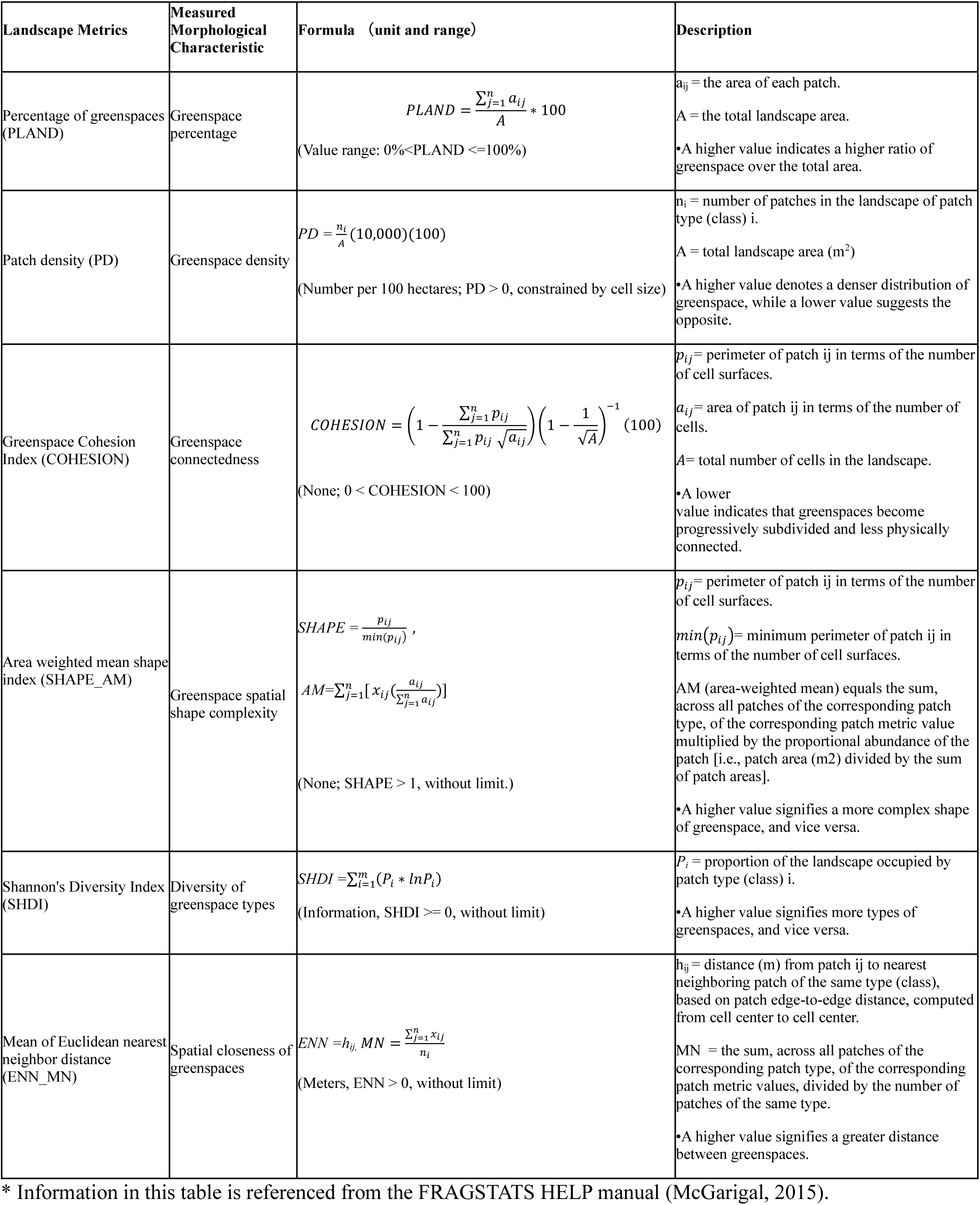
Detailed formula for calculating landscape pattern metrics.

### 2.4 Covariates

To reduce potential confounding, we adjusted all models for demographic, socioeconomic, and contextual factors consistently associated with disability risk in prior research (Dunlop et al., 2007; H. Wang & Tassinary, 2023; Wheaton & Crimmins, 2016). Because determinants of disability vary across the life course, covariate specifications were tailored to each age group while maintaining a consistent conceptual framework. Age composition was incorporated to account for underlying demographic structure within tracts, including the proportion of residents aged 65 years and older in the older adult models and the proportion of the relevant age group in adult and child models, given the strong association between aging and functional limitation(Freedman et al., 2002; Wheaton & Crimmins, 2016). Gender composition was represented by the percentage of females in each age-specific population, including female older adults, female adults aged 18–64, and female children, reflecting documented sex differences in disability prevalence. Racial composition was operationalized as the percentage of White residents to account for well-established racial and ethnic disparities in disability patterns (Dunlop et al., 2007). Educational attainment, measured as the percentage of individuals with a high school degree or less, was included across age-group models because lower educational attainment is consistently associated with increased disability risk and reduced health resilience (Mandemakers & Monden, 2010; McFarland et al., 2017).

Socioeconomic conditions were captured primarily through median household income in all models and, for older adults, the unemployment rate, because economic disadvantage is strongly associated with both disability risk and unequal neighborhood environmental conditions, including the availability and quality of greenspace. Adjusting for these variables was therefore important to reduce socioeconomic confounding in the estimated associations between greenspace morphology and disability prevalence (Smythe & Kuper, 2024). Marital status was included in the adult and older adult models because evidence suggests that marriage has social and economic protective effects among vulnerable populations (Carr et al., 2019). For children, additional household-context variables, including the percentages of households with children and single-parent households, were incorporated to reflect family-structural influences on child health outcomes (Amato, 2005; McLanahan & Sandefur, 2009). Urbanicity was represented by population density, acknowledging the influence of spatial context on environmental exposure, service accessibility, and health outcomes (Wong et al., 2023). Because the dependent variables were age-specific disability prevalence percentages rather than counts, population size was included as a covariate to account for variation in the population base across census tracts, rather than as an offset. To isolate the influence of greenspace morphology beyond overall greenspace quantity, the greenspace percentage was controlled for in the models, consistent with prior morphology–health research (H. Wang & Tassinary, 2019, 2024). Socio-demographic covariates were obtained from the 2023 American Community Survey using the *tidycensus* package in R (R Core Team, 2021; Walker, 2024).

### 2.5. Statistical analysis

We summarized age-specific disability prevalence, greenspace morphology metrics, and socio-demographic characteristics using descriptive statistics for children (5–17 years), working-age adults (18–64 years), and older adults (65 years and older), recognizing that disability patterns and neighborhood composition vary across life stages. Pairwise correlation matrices were examined within each age group to assess relationships among morphology indicators and covariates. Correlation analyses revealed moderate-to-high interdependence among several landscape metrics, reflecting the inherent mathematical relationships among morphology measures. Including highly correlated morphology metrics simultaneously may destabilize coefficient estimates and make the interpretation of results conceptually challenging; therefore, we estimated a series of models in which each greenspace morphology metric was entered separately while controlling for overall greenspace.

We assessed spatial dependence of the residual using Moran’s I and observed significant spatial autocorrelation (p < 0.001), indicating clustering of similar disability patterns across neighboring census tracts. ignoring spatial dependence can produce biased or inefficient parameter estimates and lead to incorrect statistical inference (Anselin, 1995). We therefore conducted Lagrange Multiplier spatial regression diagnostic tests and compared Spatial Error Models and Spatial Lag Models using model diagnostics and Akaike Information Criterion (AIC) values (Appendix A Table 1). Based on these comparisons, Spatial Lag Models generally demonstrated superior model performance and were therefore selected for the primary analyses. Spatial Lag Models explicitly account for spatial dependence by incorporating spatially lagged dependent variables, making them appropriate when outcomes in neighboring areas may influence one another (Bivand et al., 2013; Kissling & Carl, 2008). Spatial relationships among census tracts were defined using a row-standardized queen contiguity weights matrix, in which tracts sharing a common boundary or vertex were considered neighbors. Although count-based models were considered, negative binomial specifications are not supported in standard spatial lag implementations in R; thus, disability prevalence was used as the outcome variable instead of raw counts. Additional corrections such as county-clustered or bootstrapped standard errors were not applied, as they would be conceptually redundant within a spatial lag modeling framework.

For the children’s subgroup, we observed a significant right-skewed distribution in children’s prevalence data, a logarithmic transformation was applied to improve symmetry and model stability (Osborne, 2010). To enhance numerical stability and facilitate interpretation, all continuous independent variables were standardized prior to estimation, placing predictors on a comparable scale and reducing computational issues arising from differences in measurement units (Gelman, 2008). Multicollinearity was evaluated using variance inflation factors, and were found all below 4, indicating minimum collinearity (O’brien, 2007). Model fit was assessed using Akaike Information Criterion (AIC) (Akaike, 1974), and residual Moran’s I statistics were examined to confirm that spatial dependence had been adequately addressed within the analytical framework. All analyses were conducted in R (version 4.5.1, R Core Team, 2021) using the *spdep*, *sf*, and *car* packages (Bivand et al., 2013; Fox & Weisberg, 2018; Pebesma, 2018).

## 3. Results

3.1. **Descriptive statistics**

We analyzed 60,479 census tracts for the working-age adult group, 60,479 for the older adult group, and 25,230 for children. Descriptive statistics for disability prevalence, greenspace morphology, and socio-demographic characteristics are presented in Table 1a–c, and their spatial distributions are illustrated in Figure 1 (a-j). Disability prevalence varied substantially across age groups, with older adults exhibiting the highest mean prevalence (33.83%, SD = 13.46), followed by working-age adults (11.32%, SD = 6.65), whereas children (5–17 years) showed markedly lower rates (0.76%, SD = 0.94).

**Figure 1.**
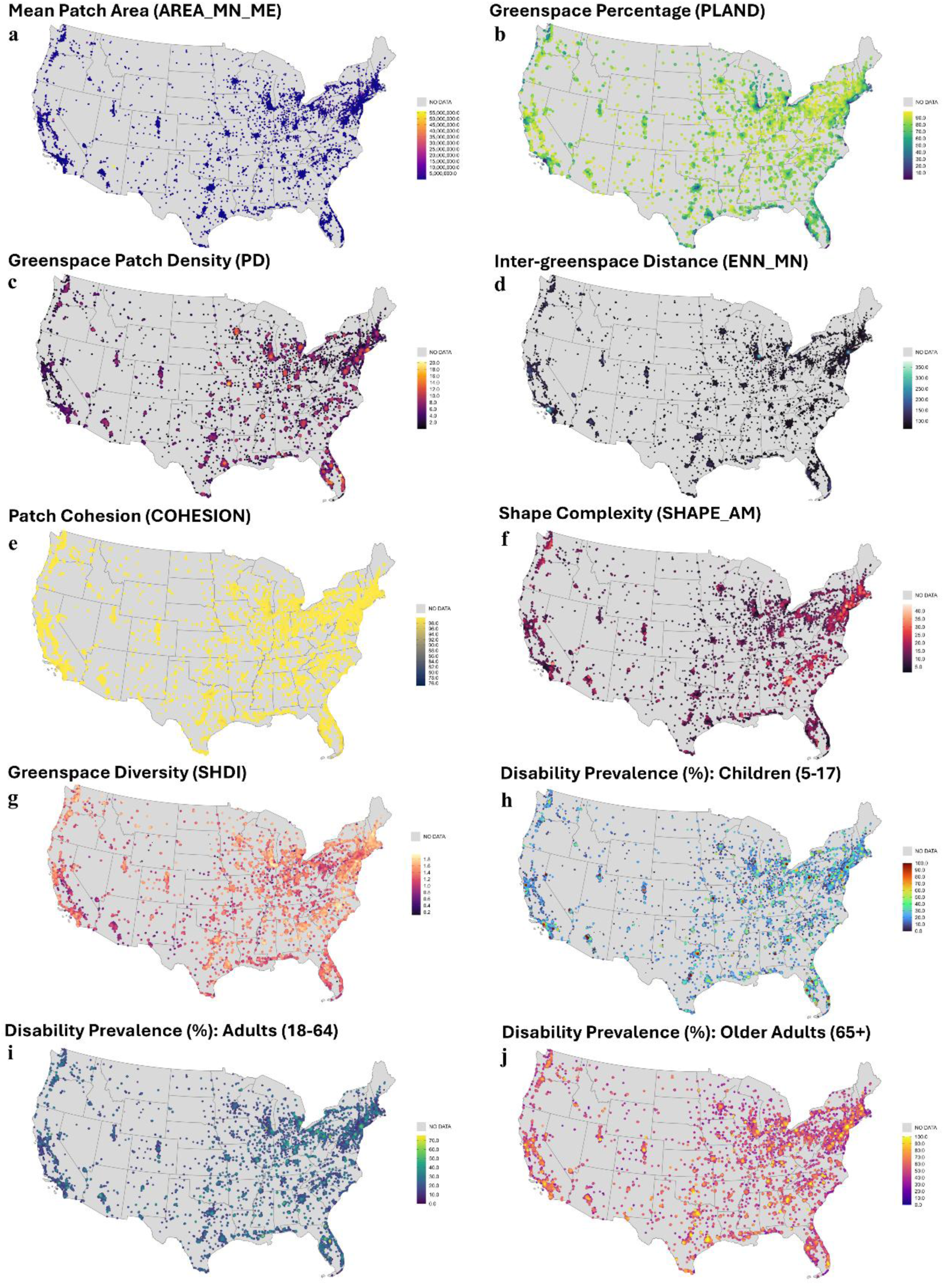
(a-j). Spatial distribution of greenspace morphology and disability prevalence in study area. Morphology indicators include: (a). mean patch area (AREA_MN), (b). percentage of greenspace (PLAND), (c). patch density (PD), (d). mean nearest neighbor distance (ENN_MN), (e). patch cohesion (COHESION), (f). shape complexity (SHAPE_AM), and (g). greenspace diversity (SHDI). Disability prevalence is shown for (h). children (5–17 years), (i). adults (18–64 years), and (j). older adults (65+ years). Bubble size and color intensity correspond to tract-level values.

Pairwise correlation matrices for each age group are presented in Tables 2–4. Across all samples, greenspace morphology metrics exhibited moderate-to-strong intercorrelations, particularly between greenspace percentage and connectedness, shape complexity, inter-greenspace distance, and diversity (|r| often > 0.5), reflecting the inherent mathematical and spatial relationships among landscape morphology indicators. Greenspace percentage was modestly and negatively correlated with disability prevalence among working-age adults (r = −0.233) and children (r = −0.090), whereas correlations were comparatively weaker among older adults (r = −0.048). Inter-greenspace distance generally showed positive correlations with disability prevalence across age groups, while connectedness and diversity tended to exhibit inverse correlations. Socio-demographic variables, particularly median household income, marital status, and educational attainment, demonstrated comparatively stronger correlations with disability prevalence and were also significantly associated with several greenspace morphology metrics across age groups, supporting their treatment as important socioeconomic confounders in the subsequent models. The observed intercorrelations among morphology metrics further supported the decision to estimate separate models for each morphology indicator rather than including them simultaneously within a single regression framework.

**Table 2:** Descriptive statistics for disability prevalence, greenspace morphology, and socio-demographic characteristics among older adults (65+).

| Variable | Mean | SD | Min | Max |
| --- | --- | --- | --- | --- |
| **Disability prevalence (%)** |  |  |  |  |
| Older adults (65+) with disability (%) | 33.825 | 13.463 | 0.000 | 100.000 |
| **Greenspace morphology** |  |  |  |  |
| Greenspace percentage | 53.890 | 31.158 | 0.469 | 99.951 |
| Greenspace Patch Density | 3.718 | 3.001 | 0.001 | 20.406 |
| Greenspace mean area (meter) | 172.786 | 1,285.202 | 0.583 | 140,233.183 |
| Greenspace connectedness | 97.814 | 3.271 | 75.134 | 100.000 |
| Greenspace shape complexity | 9.311 | 5.709 | 1.567 | 69.479 |
| Inter-greenspace distance | 95.908 | 29.585 | 59.970 | 642.679 |
| Greenspace diversity | 0.795 | 0.592 | 0.000 | 2.108 |
| **Social-demographic factors** |  |  |  |  |
| Older Adults aged 65+ (%) | 17.633 | 8.477 | 0.030 | 92.290 |
| Female (%) | 50.864 | 4.307 | 21.240 | 90.390 |
| White (%) | 63.193 | 27.057 | 0.000 | 100.000 |
| High school degree or less (%) | 11.200 | 9.652 | 0.000 | 80.790 |
| Married population (%) | 46.940 | 13.929 | 0.000 | 93.060 |
| Unemployment rate (%) | 3.426 | 2.644 | 0.000 | 45.240 |
| Median income (\$) | 84,689.981 | 40,863.984 | 2,499.000 | 250,001.000 |
| Population size | 3,911.955 | 1,670.875 | 60.000 | 24,659.000 |
| Population density (urbanicity) | 3619.068 | 6895.778 | 425.100 | 241600.800 |

**Table 3.** Descriptive statistics for disability prevalence, greenspace morphology, and socio-demographic characteristics among working-age adults (18–64).

| Variable | Mean | SD | Min | Max |
| --- | --- | --- | --- | --- |
| **Disability prevalence (%)** |  |  |  |  |
| Adults (18–64) with disability (%) | 11.318 | 6.646 | 0.000 | 79.590 |
| **Greenspace morphology** |  |  |  |  |
| Greenspace percentage | 53.890 | 31.158 | 0.469 | 99.951 |
| Greenspace Patch Density | 3.718 | 3.001 | 0.001 | 20.406 |
| Greenspace mean area (meter) | 172.786 | 1,285.202 | 0.583 | 140,233.183 |
| Greenspace connectedness | 97.814 | 3.271 | 75.134 | 100.000 |
| Greenspace shape complexity | 9.311 | 5.709 | 1.567 | 69.479 |
| Inter-greenspace distance | 95.908 | 29.585 | 59.970 | 642.679 |
| Greenspace diversity | 0.795 | 0.592 | 0.000 | 2.108 |
| **Social-demographic factors** |  |  |  |  |
| Adults aged 18–64 (%) | 60.685 | 8.115 | 6.630 | 99.590 |
| Female (%) | 50.864 | 4.307 | 21.240 | 90.390 |
| White (%) | 63.193 | 27.057 | 0.000 | 100.000 |
| High school degree or less (%) | 11.200 | 9.652 | 0.000 | 80.790 |
| Married population (%) | 46.940 | 13.929 | 0.000 | 93.060 |
| Median income (\$) | 84,689.981 | 40,863.984 | 2,499.000 | 250,001.000 |
| Population size | 3,911.955 | 1,670.875 | 60.000 | 24,659.000 |
| Population density (urbanicity) | 3619.068 | 6895.778 | 425.100 | 241600.800 |

**Table 2:** Descriptive statistics for disability prevalence, greenspace morphology, and socio-demographic characteristics among children (5–17).

| Variable | Mean | SD | Min | Max |
| --- | --- | --- | --- | --- |
| **Disability prevalence** |  |  |  |  |
| Children (5–17) with disability (%) | 0.755 | 0.944 | -3.291 | 4.605 |
| **Greenspace morphology** |  |  |  |  |
| Greenspace percentage | 58.792 | 30.739 | 0.470 | 99.951 |
| Greenspace Patch Density | 3.422 | 2.972 | 0.001 | 18.654 |
| Greenspace mean area (meter) | 204.219 | 1,307.266 | 0.584 | 140,233.183 |
| Greenspace connectedness | 98.118 | 3.145 | 76.632 | 100.000 |
| Greenspace shape complexity | 9.756 | 5.669 | 1.567 | 56.264 |
| Inter-greenspace distance | 93.452 | 28.475 | 59.980 | 377.270 |
| Greenspace diversity | 0.834 | 0.588 | 0.000 | 2.108 |
| **Social-demographic factors** |  |  |  |  |
| Children aged 5–17 (%) | 22.978 | 6.088 | 0.500 | 64.350 |
| Female children (5–17) (%) | 48.540 | 11.043 | 0.000 | 100.000 |
| White population (%) | 64.204 | 27.174 | 0.000 | 100.000 |
| High school degree or less (%) | 11.981 | 9.880 | 0.000 | 80.790 |
| Median income (\$) | 81,607.589 | 38,977.184 | 2,499.000 | 250,001.000 |
| Single-parent households (%) | 14.932 | 10.931 | 0.000 | 100.000 |
| Households with children (%) | 32.105 | 10.344 | 0.605 | 91.623 |
| Population size | 4,101.988 | 1,690.131 | 280.000 | 24,659.000 |
| Population density (urbanicity) | 2953.300 | 5228.435 | 425.300 | 95785.500 |

**Table 2.** Correlation matrix of greenspace, adult disability outcome, and socio-demographic variables: Adult (18-64)

| Variable | 1 | 2 | 3 | 4 | 5 | 6 | 7 | 8 | 9 | 10 | 11 | 12 | 13 | 14 | 15 | 16 |
| --- | --- | --- | --- | --- | --- | --- | --- | --- | --- | --- | --- | --- | --- | --- | --- | --- |
| 1 Greenspace percentage | 1.000 |  |  |  |  |  |  |  |  |  |  |  |  |  |  |  |
| 2 Greenspace Patch Density | 0.577*** | 1.000 |  |  |  |  |  |  |  |  |  |  |  |  |  |  |
| 3 Greenspace mean area | 0.175*** | 0.153*** | 1.000 |  |  |  |  |  |  |  |  |  |  |  |  |  |
| 4 Greenspace connectedness | -0.764*** | -0.299*** | -0.087*** | 1.000 |  |  |  |  |  |  |  |  |  |  |  |  |
| 5 Greenspace shape complexity | -0.613*** | -0.315*** | -0.013*** | 0.594*** | 1.000 |  |  |  |  |  |  |  |  |  |  |  |
| 6 Inter-greenspace distance | 0.704*** | 0.010* | 0.094*** | -0.760*** | 0.530*** | 1.000 |  |  |  |  |  |  |  |  |  |  |
| 7 Greenspace diversity | -0.681*** | -0.399*** | -0.088*** | -0.535*** | -0.551*** | -0.548*** | 1.000 |  |  |  |  |  |  |  |  |  |
| 8 Adult disability prevalence (%) | -0.233*** | -0.157*** | -0.075*** | -0.107*** | -0.007 | 0.156*** | -0.112*** | 1.000 |  |  |  |  |  |  |  |  |
| 9 Adults aged 18–64 (%) | 0.257*** | 0.137*** | 0.063*** | -0.211*** | -0.178*** | -0.225*** | 0.229*** | -0.089*** | 1.000 |  |  |  |  |  |  |  |
| 10 White (%) | -0.480*** | -0.256*** | -0.046*** | 0.386*** | 0.337*** | -0.424*** | 0.416*** | -0.039*** | -0.215*** | 1.000 |  |  |  |  |  |  |
| 11 High school degree or less (%) | 0.132*** | -0.078*** | -0.020*** | -0.218*** | -0.180*** | 0.270*** | -0.212*** | 0.264*** | 0.041*** | -0.516*** | 1.000 |  |  |  |  |  |
| 12 Median income (\$) | 0.131*** | 0.111*** | 0.075*** | 0.033*** | 0.139*** | 0.057*** | -0.005 | -0.547*** | -0.014*** | 0.199*** | -0.456*** | 1.000 | | | | |
| 13 Female (%) | 0.063*** | 0.096*** | 0.022*** | -0.019*** | -0.038*** | -0.002 | -0.016*** | 0.049*** | -0.115*** | -0.120*** | -0.043*** | -0.091*** | 1.000 |  |  |  |
| 14 Population density (urbanicity) | 0.428*** | 0.028*** | 0.052*** | -0.333*** | -0.282*** | 0.465*** | -0.380*** | -0.106*** | 0.233*** | -0.264*** | 0.127*** | 0.013** | 0.038*** | 1.000 |  |  |
| 15 Married population (%) | -0.306*** | -0.179*** | -0.032*** | 0.298*** | 0.337*** | -0.250*** | 0.334*** | -0.335*** | -0.420*** | 0.526*** | -0.393*** | 0.569*** | -0.139*** | -0.233*** | 1.000 |  |
| 16 Population size | 0.028*** | 0.081*** | 0.083*** | 0.039*** | 0.101*** | 0.004 | 0.064*** | -0.223*** | 0.039*** | -0.043*** | -0.055*** | 0.233*** | 0.002 | 0.047*** | 0.168*** | 1.000 |
| *** Statistically significant at the 0.001 level. |  |  |  |  |  |  |  |  |  |  |  |  |  |  |  |  |
| ** Statistically significant at the 0.01 level. |  |  |  |  |  |  |  |  |  |  |  |  |  |  |  |  |
| * Statistically significant at the 0.05 level. |  |  |  |  |  |  |  |  |  |  |  |  |  |  |  |  |

**Table 3.** Correlation matrix of greenspace, older-adults disability outcome, and socio-demographic variables: Older Adults (65+)

| Variable | 1 | 2 | 3 | 4 | 5 | 6 | 7 | 8 | 9 | 10 | 11 | 12 | 13 | 14 | 15 | 16 | 17 |
| --- | --- | --- | --- | --- | --- | --- | --- | --- | --- | --- | --- | --- | --- | --- | --- | --- | --- |
| 1 Greenspace percentage | 1.000 |  |  |  |  |  |  |  |  |  |  |  |  |  |  |  |  |
| 2 Greenspace Patch Density | 0.577*** | 1.000 |  |  |  |  |  |  |  |  |  |  |  |  |  |  |  |
| 3 Greenspace mean area | 0.175*** | 0.153*** | 1.000 |  |  |  |  |  |  |  |  |  |  |  |  |  |  |
| 4 Greenspace connectedness | -0.764*** | -0.299*** | -0.087*** | 1.000 |  |  |  |  |  |  |  |  |  |  |  |  |  |
| 5 Greenspace shape complexity | -0.613*** | -0.315*** | -0.013*** | 0.594*** | 1.000 |  |  |  |  |  |  |  |  |  |  |  |  |
| 6 Inter-greenspace distance | 0.704*** | 0.010* | 0.094*** | -0.760*** | 0.530*** | 1.000 |  |  |  |  |  |  |  |  |  |  |  |
| 7 Greenspace diversity | -0.681*** | -0.399*** | -0.088*** | -0.535*** | -0.551*** | -0.548*** | 1.000 |  |  |  |  |  |  |  |  |  |  |
| 8 Older-adult disability prevalence (%) | -0.048*** | -0.094*** | -0.053*** | -0.029*** | -0.086*** | 0.038*** | -0.036*** | 1.000 |  |  |  |  |  |  |  |  |  |
| 9 Older adults aged 65+ (%) | -0.158*** | -0.140*** | -0.064*** | 0.145*** | 0.108*** | -0.167*** | 0.194*** | -0.088*** | 1.000 |  |  |  |  |  |  |  |  |
| 10 Female (%) | 0.063*** | 0.096*** | 0.022*** | -0.019*** | -0.038*** | 0.002 | -0.016*** | 0.060*** | 0.080*** | 1.000 |  |  |  |  |  |  |  |
| 11 White (%) | -0.480*** | -0.256*** | -0.046*** | 0.386*** | 0.337*** | -0.424*** | 0.416*** | -0.186*** | 0.360*** | -0.120*** | 1.000 |  |  |  |  |  |  |
| 12 High school degree or less (%) | 0.132*** | -0.078*** | -0.020*** | -0.218*** | -0.180*** | 0.270*** | -0.212*** | 0.339*** | -0.243*** | -0.043*** | -0.516*** | 1.000 |  |  |  |  |  |
| 13 Married population (%) | -0.306*** | -0.179*** | -0.032*** | 0.298*** | 0.337*** | -0.250*** | 0.334*** | -0.327*** | 0.315*** | -0.139*** | 0.526*** | -0.393*** | 1.000 |  |  |  |  |
| 14 Unemployment rate (%) | 0.208*** | 0.075*** | 0.022*** | -0.189*** | -0.173*** | 0.200*** | -0.222*** | 0.157*** | -0.229*** | 0.044*** | -0.392*** | 0.278*** | -0.386*** | 1.000 |  |  |  |
| 15 Median income (\$) | 0.131*** | 0.111*** | 0.075*** | 0.033*** | 0.139*** | 0.057*** | -0.005 | -0.410*** | 0.023*** | -0.091*** | 0.199*** | -0.456*** | 0.569*** | -0.228*** | 1.000 | | |
| 16 Population size | 0.028*** | 0.081*** | 0.083*** | 0.039*** | 0.101*** | 0.004 | 0.064*** | -0.090*** | -0.203*** | 0.002 | -0.043*** | -0.055*** | 0.168*** | -0.039*** | 0.233*** | 1.000 |  |
| 17 Population density (urbanicity) | 0.428*** | 0.028*** | 0.052*** | -0.333*** | -0.282*** | 0.465*** | -0.380*** | 0.036*** | -0.121*** | 0.038*** | -0.264*** | 0.127*** | -0.233*** | 0.150*** | 0.013** | 0.047*** | 1.000 |
| *** Statistically significant at the 0.001 level. |  |  |  |  |  |  |  |  |  |  |  |  |  |  |  |  |  |
| ** Statistically significant at the 0.01 level. |  |  |  |  |  |  |  |  |  |  |  |  |  |  |  |  |  |
| * Statistically significant at the 0.05 level. |  |  |  |  |  |  |  |  |  |  |  |  |  |  |  |  |  |

**Table 4.** Correlation matrix of greenspace, Children’s disability outcome, and socio-demographic variables: Children (5-7)

| Variable | 1 | 2 | 3 | 4 | 5 | 6 | 7 | 8 | 9 | 10 | 11 | 12 | 13 | 14 | 15 | 16 | 17 |
| --- | --- | --- | --- | --- | --- | --- | --- | --- | --- | --- | --- | --- | --- | --- | --- | --- | --- |
| 1 Greenspace percentage | 1.000 |  |  |  |  |  |  |  |  |  |  |  |  |  |  |  |  |
| 2 Greenspace Patch Density | 0.650*** | 1.000 |  |  |  |  |  |  |  |  |  |  |  |  |  |  |  |
| 3 Greenspace mean area | 0.182*** | -0.165*** | 1.000 |  |  |  |  |  |  |  |  |  |  |  |  |  |  |
| 4 Greenspace connectedness | -0.763*** | -0.343*** | 0.090*** | 1.000 |  |  |  |  |  |  |  |  |  |  |  |  |  |
| 5 Greenspace shape complexity | -0.599*** | -0.357*** | 0.017** | 0.576*** | 1.000 |  |  |  |  |  |  |  |  |  |  |  |  |
| 6 Inter-greenspace distance | 0.695*** | -0.080*** | 0.103*** | -0.778*** | 0.510*** | 1.000 |  |  |  |  |  |  |  |  |  |  |  |
| 7 Greenspace diversity | -0.664*** | -0.435*** | 0.089*** | 0.523*** | 0.537*** | 0.531*** | 1.000 |  |  |  |  |  |  |  |  |  |  |
| 8 Children disability prevalence (%) | -0.090*** | -0.056*** | -0.021*** | -0.074*** | 0.092*** | 0.040*** | -0.063*** | 1.000 |  |  |  |  |  |  |  |  |  |
| 9 Children aged 5–17 (%) | 0.011 | 0.050*** | -0.015* | -0.005 | 0.012 | -0.015* | -0.064*** | -0.222*** | 1.000 |  |  |  |  |  |  |  |  |
| 10 White population (%) | -0.532*** | -0.305*** | 0.050*** | 0.423*** | 0.366*** | 0.463*** | 0.450*** | -0.075*** | -0.239*** | 1.000 |  |  |  |  |  |  |  |
| 11 High school degree or less (%) | -0.198*** | -0.058*** | 0.015* | -0.285*** | -0.224*** | -0.354*** | -0.270*** | 0.042*** | 0.258*** | -0.533*** | 1.000 |  |  |  |  |  |  |
| 12 Median income (\$) | -0.125*** | 0.161*** | -0.085*** | 0.042*** | 0.148*** | -0.024*** | 0.009 | -0.158*** | -0.012 | 0.175*** | -0.458*** | 1.000 | | | | | |
| 13 Female children (5–17) (%) | -0.005 | 0.008 | 0.006 | 0.004 | -0.000 | 0.005 | -0.002 | -0.022*** | 0.017** | -0.009 | -0.006 | 0.006 | 1.000 |  |  |  |  |
| 14 Single-parent households (%) | -0.180*** | 0.157*** | -0.037*** | -0.172*** | -0.230*** | -0.123*** | -0.226*** | 0.098*** | 0.343*** | -0.459*** | 0.396*** | -0.513*** | 0.017** | 1.000 |  |  |  |
| 15 Households with children (%) | -0.079*** | 0.061*** | -0.042*** | -0.059*** | 0.006 | -0.126*** | -0.100*** | -0.217*** | 0.780*** | -0.310*** | 0.297*** | 0.198*** | 0.011 | 0.217*** | 1.000 |  |  |
| 16 Population density (urbanicity) | -0.448*** | 0.110*** | -0.061*** | -0.337*** | -0.288*** | -0.428*** | -0.400*** | 0.074*** | -0.026*** | -0.350*** | 0.225*** | -0.057*** | 0.007 | 0.156*** | 0.001 | 1.000 |  |
| 17 Population size | -0.082*** | 0.107*** | -0.096*** | 0.006 | 0.079*** | -0.043*** | 0.035*** | -0.164*** | 0.181*** | -0.078*** | -0.056*** | 0.276*** | 0.012* | -0.109*** | 0.299*** | 0.067*** | 1.000 |

\*\*\* Statistically significant at the 0.001 level.
\*\* Statistically significant at the 0.01 level.
\* Statistically significant at the 0.05 level.

### 3.2. Associations between greenspace morphology and disability prevalence

Tables 5–7 present the spatial lag model estimates for the associations between greenspace morphology metrics and disability prevalence across the three age groups. Overall, lower disability prevalence tended to occur in neighborhoods with higher greenspace percentage, density, connectedness, complex shape, greater diversity, and more clustered greenspace distributions, although the strength of these relationships varied by age group.

**Table 5.**
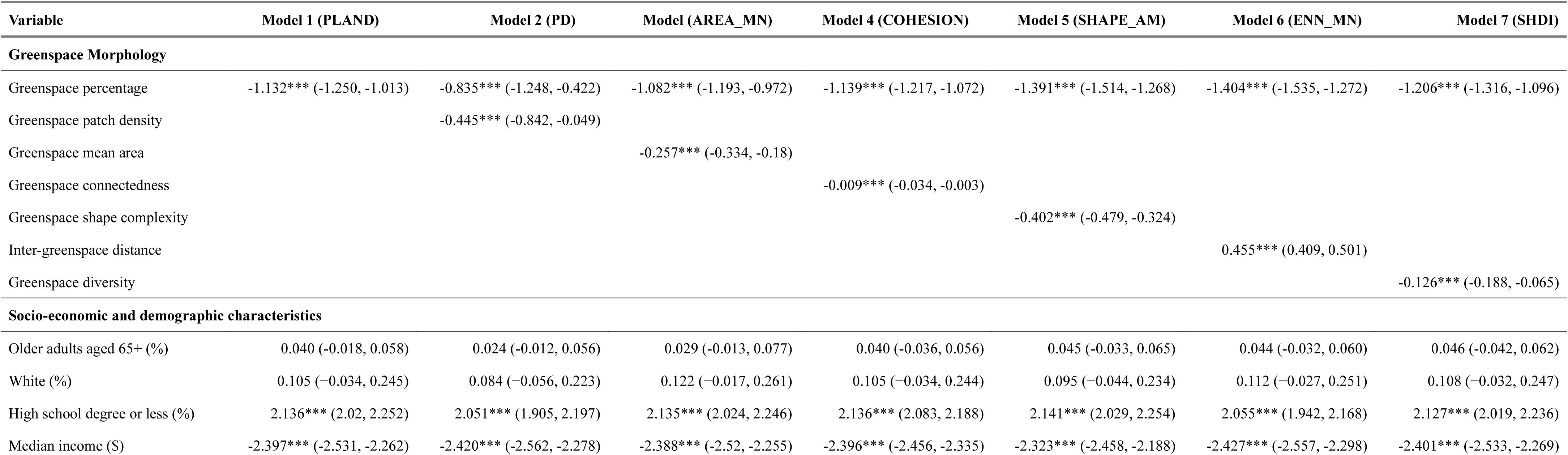
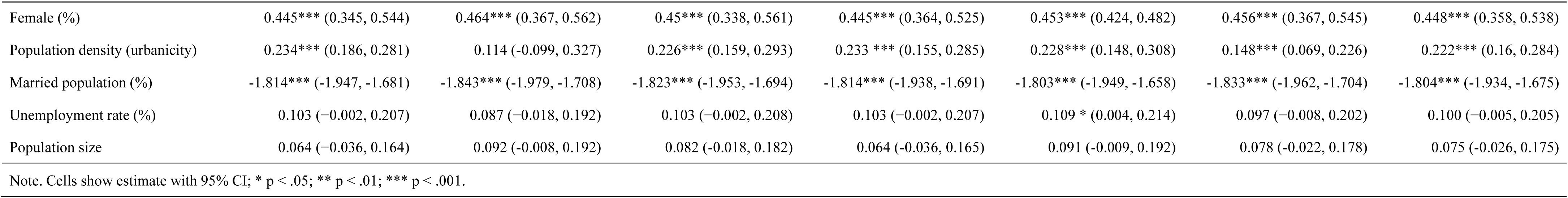
Regression coefficients of greenspace and prevalence of disability relationship among older adults.

| Variable | Model 1 (PLAND) | Model 2 (PD) | Model (AREA_MN) | Model 4 (COHESION) | Model 5 (SHAPE_AM) | Model 6 (ENN_MN) | Model 7 (SHDI) |
| --- | --- | --- | --- | --- | --- | --- | --- |
| Greenspace Morphology |  |  |  |  |  |  |  |
| Greenspace percentage | -1.132*** (-1.250, -1.013) | -0.835*** (-1.248, -0.422) | -1.082*** (-1.193, -0.972) | -1.139*** (-1.217, -1.072) | -1.391*** (-1.514, -1.268) | -1.404*** (-1.535, -1.272) | -1.206*** (-1.316, -1.096) |
| Greenspace patch density |  | -0.445*** (-0.842, -0.049) |  |  |  |  |  |
| Greenspace mean area |  |  | -0.257*** (-0.334, -0.18) |  |  |  |  |
| Greenspace connectedness |  |  |  | -0.009*** (-0.034, -0.003) |  |  |  |
| Greenspace shape complexity |  |  |  |  | -0.402*** (-0.479, -0.324) |  |  |
| Inter-greenspace distance |  |  |  |  |  | 0.455*** (0.409, 0.501) |  |
| Greenspace diversity |  |  |  |  |  |  | -0.126*** (-0.188, -0.065) |
| Socio-economic and demographic characteristics |  |  |  |  |  |  |  |
| Older adults aged 65+ (%) | 0.040 (-0.018, 0.058) | 0.024 (-0.012, 0.056) | 0.029 (-0.013, 0.077) | 0.040 (-0.036, 0.056) | 0.045 (-0.033, 0.065) | 0.044 (-0.032, 0.060) | 0.046 (-0.042, 0.062) |
| White (%) | 0.105 (-0.034, 0.245) | 0.084 (-0.056, 0.223) | 0.122 (-0.017, 0.261) | 0.105 (-0.034, 0.244) | 0.095 (-0.044, 0.234) | 0.112 (-0.027, 0.251) | 0.108 (-0.032, 0.247) |
| High school degree or less (%) | 2.136*** (2.02, 2.252) | 2.051*** (1.905, 2.197) | 2.135*** (2.024, 2.246) | 2.136*** (2.083, 2.188) | 2.141*** (2.029, 2.254) | 2.055*** (1.942, 2.168) | 2.127*** (2.019, 2.236) |
| Median income (\$) | -2.397*** (-2.531, -2.262) | -2.420*** (-2.562, -2.278) | -2.388*** (-2.52, -2.255) | -2.396*** (-2.456, -2.335) | -2.323*** (-2.458, -2.188) | -2.427*** (-2.557, -2.298) | -2.401*** (-2.533, -2.269) |
| Female (%) | 0.445*** (0.345, 0.544) | 0.464*** (0.367, 0.562) | 0.45*** (0.338, 0.561) | 0.445*** (0.364, 0.525) | 0.453*** (0.424, 0.482) | 0.456*** (0.367, 0.545) | 0.448*** (0.358, 0.538) |
| Population density (urbanicity) | 0.234*** (0.186, 0.281) | 0.114 (-0.099, 0.327) | 0.226*** (0.159, 0.293) | 0.233 *** (0.155, 0.285) | 0.228*** (0.148, 0.308) | 0.148*** (0.069, 0.226) | 0.222*** (0.16, 0.284) |
| Married population (%) | -1.814*** (-1.947, -1.681) | -1.843*** (-1.979, -1.708) | -1.823*** (-1.953, -1.694) | -1.814*** (-1.938, -1.691) | -1.803*** (-1.949, -1.658) | -1.833*** (-1.962, -1.704) | -1.804*** (-1.934, -1.675) |
| Unemployment rate (%) | 0.103 (-0.002, 0.207) | 0.087 (-0.018, 0.192) | 0.103 (-0.002, 0.208) | 0.103 (-0.002, 0.207) | 0.109 * (0.004, 0.214) | 0.097 (-0.008, 0.202) | 0.100 (-0.005, 0.205) |
| Population size | 0.064 (-0.036, 0.164) | 0.092 (-0.008, 0.192) | 0.082 (-0.018, 0.182) | 0.064 (-0.036, 0.165) | 0.091 (-0.009, 0.192) | 0.078 (-0.022, 0.178) | 0.075 (-0.026, 0.175) |
| Note. Cells show estimate with 95% CI; * p < .05; ** p < .01; *** p < .001. |  |  |  |  |  |  |  |

**Table 6.** Regression coefficients of greenspace and prevalence of disability relationship among adults.

| Variable | Model 1 (PLAND) | Model 2 (PD) | Model 3 (AREA_MN) | Model 4 (COHESION) | Model 5 (SHAPE_AM) | Model 6 (ENN_MN) | Model 7 (SHDI) |
| --- | --- | --- | --- | --- | --- | --- | --- |
| **Greenspace Morphology** |  |  |  |  |  |  |  |
| Greenspace percentage | -0.804*** (-0.858, -0.750) | -0.812*** (-0.875, -0.749) | -0.799*** (-0.849, -0.750) | -0.701*** (-0.777, -0.625) | -0.884*** (-0.944, -0.825) | -0.715*** (-0.806, -0.624) | -0.720*** (-0.804, -0.635) |
| Greenspace patch density |  | -0.013 (-0.048, 0.022) |  |  |  |  |  |
| Greenspace mean area |  |  | -0.023* (-0.090, -0.002) |  |  |  |  |
| Greenspace connectedness |  |  |  | -0.127** (-0.243, -0.011) |  |  |  |
| Greenspace shape complexity |  |  |  |  | -0.123*** (-0.174, -0.072) |  |  |
| Inter-greenspace distance |  |  |  |  |  | 0.151** (0.059, 0.242) |  |
| Greenspace diversity |  |  |  |  |  |  | -0.146*** (-0.201, -0.091) |
| **Socio-economic and demographic characteristics** |  |  |  |  |  |  |  |
| Adults aged 18–64 (%) | -0.645*** (-0.698, -0.592) | -0.645*** (-0.696, -0.593) | -0.645*** (-0.686, -0.603) | -0.641*** (-0.688, -0.595) | -0.646*** (-0.691, -0.601) | -0.639*** (-0.687, -0.591) | -0.645*** (-0.690, -0.599) |
| White (%) | -0.484*** (-0.540, -0.427) | -0.484*** (-0.557, -0.412) | -0.485*** (-0.541, -0.429) | -0.490*** (-0.549, -0.431) | -0.482*** (-0.534, -0.429) | -0.480*** (-0.541, -0.420) | -0.480*** (-0.557, -0.402) |
| High school degree or less (%) | 0.400*** (0.349, 0.450) | 0.402*** (0.339, 0.466) | 0.400*** (0.349, 0.450) | 0.412*** (0.358, 0.465) | 0.400*** (0.355, 0.445) | 0.429*** (0.366, 0.491) | 0.411*** (0.356, 0.467) |
| Median income (\$) | -1.411*** (-1.471, -1.352) | -1.411*** (-1.47, -1.352) | -1.410*** (-1.469, -1.351) | -1.425*** (-1.485, -1.365) | -1.391*** (-1.450, -1.331) | -1.405*** (-1.465, -1.344) | -1.406*** (-1.467, -1.346) |
| Female (%) | 0.031 (-0.005, 0.067) | 0.032* (0.007, 0.056) | 0.031* (0.007, 0.058) | 0.034* (0.007, 0.061) | 0.028 (-0.005, 0.065) | 0.034* (0.008, 0.061) | 0.036* (0.008, 0.064) |
| Population density (urbanicity) | -0.145*** (-0.207, -0.083) | -0.142*** (-0.199, -0.084) | -0.146*** (-0.187, -0.105) | -0.145*** (-0.192, -0.098) | -0.147*** (-0.169, -0.126) | -0.117*** (-0.167, -0.066) | -0.132** (-0.210, -0.053) |
| Married population (%) | -1.305*** (-1.373, -1.237) | -1.304*** (-1.376, -1.232) | -1.306*** (-1.367, -1.245) | -1.299*** (-1.365, -1.234) | -1.302*** (-1.367, -1.237) | -1.296*** (-1.362, -1.229) | -1.319*** (-1.394, -1.244) |
| Unemployment rate (%) | 0.360*** (0.316, 0.404) | 0.360*** (0.303, 0.418) | 0.360*** (0.316, 0.404) | 0.360*** (0.316, 0.403) | 0.362*** (0.317, 0.407) | 0.362*** (0.317, 0.407) | 0.364*** (0.321, 0.408) |
| Population size | -0.344*** (-0.388, -0.301) | -0.345*** (-0.388, -0.303) | -0.343*** (-0.383, -0.302) | -0.348*** (-0.392, -0.305) | -0.337*** (-0.379, -0.294) | -0.349*** (-0.392, -0.307) | -0.356*** (-0.412, -0.299) |
| Note. Cells show estimate with 95% CI; * p < .05; ** p < .01; *** p < .001. |  |  |  |  |  |  |  |

**Table 7.**
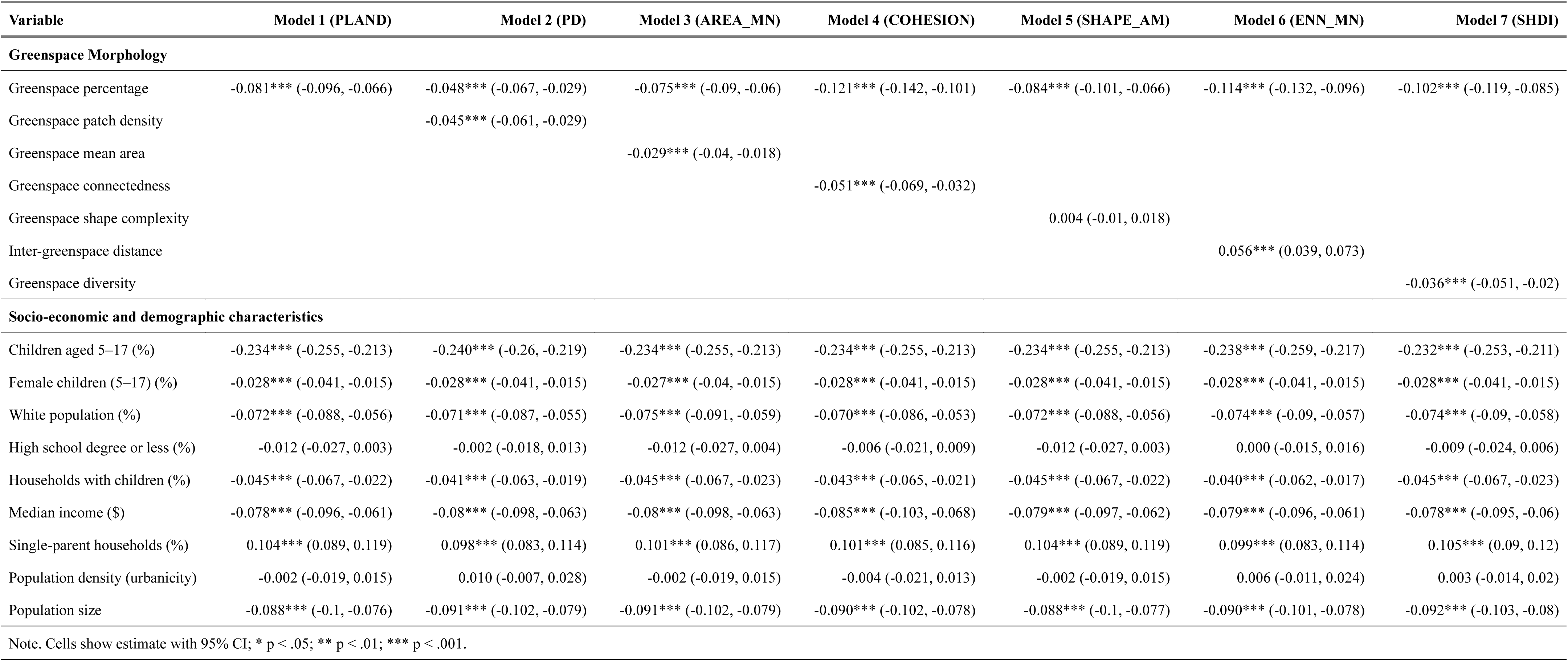
Regression coefficients of greenspace and prevalence of disability relationship among children.

| Variable | Model 1 (PLAND) | Model 2 (PD) | Model 3 (AREA_MN) | Model 4 (COHESION) | Model 5 (SHAPE_AM) | Model 6 (ENN_MN) | Model 7 (SHDI) |
| --- | --- | --- | --- | --- | --- | --- | --- |
| **Greenspace Morphology** |  |  |  |  |  |  |  |
| Greenspace percentage | -0.081*** (-0.096, -0.066) | -0.048*** (-0.067, -0.029) | -0.075*** (-0.09, -0.06) | -0.121*** (-0.142, -0.101) | -0.084*** (-0.101, -0.066) | -0.114*** (-0.132, -0.096) | -0.102*** (-0.119, -0.085) |
| Greenspace patch density |  | -0.045*** (-0.061, -0.029) |  |  |  |  |  |
| Greenspace mean area |  |  | -0.029*** (-0.04, -0.018) |  |  |  |  |
| Greenspace connectedness |  |  |  | -0.051*** (-0.069, -0.032) |  |  |  |
| Greenspace shape complexity |  |  |  |  | 0.004 (-0.01, 0.018) |  |  |
| Inter-greenspace distance |  |  |  |  |  | 0.056*** (0.039, 0.073) |  |
| Greenspace diversity |  |  |  |  |  |  | -0.036*** (-0.051, -0.02) |
| **Socio-economic and demographic characteristics** |  |  |  |  |  |  |  |
| Children aged 5–17 (%) | -0.234*** (-0.255, -0.213) | -0.240*** (-0.26, -0.219) | -0.234*** (-0.255, -0.213) | -0.234*** (-0.255, -0.213) | -0.234*** (-0.255, -0.213) | -0.238*** (-0.259, -0.217) | -0.232*** (-0.253, -0.211) |
| Female children (5–17) (%) | -0.028*** (-0.041, -0.015) | -0.028*** (-0.041, -0.015) | -0.027*** (-0.04, -0.015) | -0.028*** (-0.041, -0.015) | -0.028*** (-0.041, -0.015) | -0.028*** (-0.041, -0.015) | -0.028*** (-0.041, -0.015) |
| White population (%) | -0.072*** (-0.088, -0.056) | -0.071*** (-0.087, -0.055) | -0.075*** (-0.091, -0.059) | -0.070*** (-0.086, -0.053) | -0.072*** (-0.088, -0.056) | -0.074*** (-0.09, -0.057) | -0.074*** (-0.09, -0.058) |
| High school degree or less (%) | -0.012 (-0.027, 0.003) | -0.002 (-0.018, 0.013) | -0.012 (-0.027, 0.004) | -0.006 (-0.021, 0.009) | -0.012 (-0.027, 0.003) | 0.000 (-0.015, 0.016) | -0.009 (-0.024, 0.006) |
| Households with children (%) | -0.045*** (-0.067, -0.022) | -0.041*** (-0.063, -0.019) | -0.045*** (-0.067, -0.023) | -0.043*** (-0.065, -0.021) | -0.045*** (-0.067, -0.022) | -0.040*** (-0.062, -0.017) | -0.045*** (-0.067, -0.023) |
| Median income (\$) | -0.078*** (-0.096, -0.061) | -0.08*** (-0.098, -0.063) | -0.08*** (-0.098, -0.063) | -0.085*** (-0.103, -0.068) | -0.079*** (-0.097, -0.062) | -0.079*** (-0.096, -0.061) | -0.078*** (-0.095, -0.06) |
| Single-parent households (%) | 0.104*** (0.089, 0.119) | 0.098*** (0.083, 0.114) | 0.101*** (0.086, 0.117) | 0.101*** (0.085, 0.116) | 0.104*** (0.089, 0.119) | 0.099*** (0.083, 0.114) | 0.105*** (0.09, 0.12) |
| Population density (urbanicity) | -0.002 (-0.019, 0.015) | 0.010 (-0.007, 0.028) | -0.002 (-0.019, 0.015) | -0.004 (-0.021, 0.013) | -0.002 (-0.019, 0.015) | 0.006 (-0.011, 0.024) | 0.003 (-0.014, 0.02) |
| Population size | -0.088*** (-0.1, -0.076) | -0.091*** (-0.102, -0.079) | -0.091*** (-0.102, -0.079) | -0.090*** (-0.102, -0.078) | -0.088*** (-0.1, -0.077) | -0.090*** (-0.101, -0.078) | -0.092*** (-0.103, -0.08) |
| Note. Cells show estimate with 95% CI; * p < .05; ** p < .01; *** p < .001. |  |  |  |  |  |  |  |

Among older adults aged 65 years and older (Table 5), the results suggested a broadly protective pattern for several greenspace morphology metrics. Across model specifications, greenspace percentage was consistently and inversely associated with disability prevalence, with coefficients ranging from −0.835 to −1.404; all corresponding 95% confidence intervals excluded zero. Beyond overall greenspace percentage, higher patch density was associated with lower disability prevalence (β = −0.445, 95% CI: −0.842 to −0.049), as was larger mean patch area (β = −0.257, 95% CI: −0.334 to −0.180). Greater connectedness also showed an inverse association with disability prevalence (β = −0.009, 95% CI: −0.034 to −0.003). In contrast, inter-greenspace distance was positively associated with disability prevalence (β = 0.455, 95% CI: 0.409 to 0.501), indicating that greater spatial separation between greenspace patches was linked to higher disability prevalence. Shape complexity was also inversely associated with disability prevalence in this group (β = −0.402, 95% CI: −0.479 to −0.324). Greenspace diversity likewise showed a significant inverse association with disability prevalence (β = −0.126, 95% CI: −0.188 to −0.065).

For working-age adults aged 18–64 years (Table 6), the general pattern was similar, although fewer morphology metrics were statistically significant. Greenspace percentage remained consistently associated with lower disability prevalence across model specifications, with coefficients ranging from −0.701 to −0.884, and all corresponding 95% confidence intervals excluding zero. Larger mean patch area was modestly associated with lower disability prevalence (β = −0.023, 95% CI: −0.090 to −0.002). Greater connectedness also showed a significant inverse association with disability prevalence (β = −0.127, 95% CI: −0.243 to −0.011). In contrast, inter-greenspace distance was positively associated with disability prevalence (β = 0.151, 95% CI: 0.059 to 0.242), indicating that greater spatial separation between greenspace patches was linked to higher disability prevalence. Greenspace diversity was also inversely associated with disability prevalence (β = −0.146, 95% CI: −0.201 to −0.091). Shape complexity likewise showed a significant inverse association (β = −0.123, 95% CI: −0.174 to −0.072). In contrast, patch density was not significantly associated with disability prevalence in this age group (β = −0.013, 95% CI: −0.048 to 0.022).

Among children aged 5–17 years (Table 7), the protective pattern was generally consistent across most morphology metrics. Greenspace percentage was inversely associated with disability prevalence in all model specifications, with coefficients ranging from −0.048 to −0.121 and all corresponding 95% confidence intervals excluding zero. Higher patch density was associated with lower disability prevalence (β = −0.045, 95% CI: −0.061 to −0.029), as were larger mean patch area (β = −0.029, 95% CI: −0.040 to −0.018) and greater connectedness (β = −0.051, 95% CI: −0.069 to −0.032). In contrast, inter-greenspace distance was positively associated with disability prevalence (β = 0.056, 95% CI: 0.039 to 0.073), indicating that greater spatial separation between greenspace patches was associated with higher disability prevalence. Greenspace diversity was associated with lower disability prevalence (β = −0.036, 95% CI: −0.051 to −0.020). Shape complexity was not significantly associated with disability prevalence in the children’s models (β = 0.004, 95% CI: −0.010 to 0.018). Socio-demographic covariates showed generally stable associations across model specifications and are reported in Tables 5–7.

## 4. Discussion

### 4.1 Summary of key contributions and findings

To the best of our knowledge, this is the first national-scale study to examine the association between greenspace morphology and disability prevalence. Additionally, this is the first study to have examined such an association across the life course, across children, working-age adults, and older adults. More importantly, we examined such association with the percentage of greenspace controlled in the statistical model, which allows us to ascertain the relationship beyond the influence of green abundance or greenness. Our findings show that, after accounting for spatial dependence and adjusting for established demographic, socioeconomic, and contextual factors known to shape disability patterns, and controlling for the percentage of greenspace, greenspace morphology explains additional variance in disability risk. Specifically, greater diversity, larger patch area, stronger connectedness, higher patch density, and shorter inter-greenspace distance were generally associated with lower disability prevalence, although the strength and consistency of these associations varied across age groups. These patterns were observed most consistently among children and older adults, whereas associations among working-age adults were more selective. More importantly, the findings indicate that several dimensions of greenspace morphology were associated with disability prevalence beyond overall vegetation coverage, underscoring the importance of how greenspace is spatially structured within urban neighborhoods.

### 4.2 Greenspace abundance and disability risk

This study builds upon existing evidence by first reaffirming the inverse relationship between greenspace abundance and disability prevalence across urban neighborhoods in the United States. In particular, the consistently negative association between greenspace percentage and disability prevalence across children, working-age adults, and older adults aligns with prior research showing that greater greenspace exposure is associated with lower disability risk or slower functional decline in different populations, including older adults with limitations in activities of daily living (Peng et al., 2020) and poststroke disability (Cao et al., 2023). We are aware of another nationwide study reported that areas with higher greenspace had higher disability rates (Wong et al., 2023). One likely reason for this difference is that the prior study included both urban and rural areas, whereas the present study focused specifically on urban census tracts. Because disability prevalence is generally higher in rural areas, and because rural–urban differences are also tied to broader socioeconomic and environmental inequalities, the prior association may partly reflect structural differences across place types rather than the specific role of greenspace alone. In the present study, we sought to reduce this source of bias by restricting the analysis to urban tracts and by adjusting for a broad set of demographic and socioeconomic covariates. While residual confounding cannot be ruled out, these design choices provide greater confidence that the observed associations reflect neighborhood greenspace conditions within urban settings rather than broader rural–urban or socioeconomic disparities.

### 4.3 Greenspace morphology and disability risk

After adjusting for greenspace percentage, thereby holding overall greenspace abundance constant, we found that multiple greenspace morphology metrics remained associated with disability outcomes across children, working-age adults, and older adults. These associations were more pervasive and consistent among older adults and children, whereas they were more selective among working-age adults. This life-course pattern suggests that greenspace morphology may be particularly salient when limited daily mobility, greater dependency, or heightened vulnerability to neighborhood conditions makes the immediate residential environment more influential (Ben-Shlomo & Kuh, 2002; Clarke et al., 2008; Markevych et al., 2017). In contrast, working-age adults may spend substantial portions of the day away from home (e.g., at workplaces), which could attenuate the influence of greenspace morphology on their disability risk.

Specifically, we found that higher greenspace density, larger patch size, greater connectedness, lower inter-greenspace distance, greater shape complexity, and higher diversity were associated with lower disability prevalence in several models and age groups, although the observed patterns varied across the life course. These findings are consistent with prior city-level evidence suggesting that greenspace morphology is meaningfully related to disability prevalence. For example, earlier work conducted in a large U.S. metropolitan context showed that larger, less fragmented, and more connected greenspace configurations were associated with lower disability prevalence, with behavioral pathways such as physical activity and mental health playing an important mediating role (Xu & Wang, 2025). Building on this earlier evidence, the present study extends the scope to a national urban system and demonstrates that morphology–disability associations are observable across diverse urban contexts and population age groups, even when overall greenspace coverage is held constant.

Additionally, the association between greenspace morphology and disability risk also aligns with some observations linking greenspace morphology to other health outcomes. Larger and more connected greenspace patches have been linked to lower risks of cardiovascular disease, obesity, pregnancy outcomes, and all-cause mortality, while more spatially clustered greenspaces have been associated with better mental health (H. Wang, Huang, et al., 2024; H. Wang & Tassinary, 2019, 2024). Morphological attributes such as connectivity and proximity have also been shown to promote leisure-time physical activity and walking behavior beyond the effects of total greenspace coverage (H. Wang & Tassinary, 2024), outcomes that are closely tied to functional capacity and disability risk. A recent systematic review also documented that morphological characteristics such as larger patch size, greater connectedness, higher aggregation, lower fragmentation, and greater compositional diversity are associated with reduced risks of cardiovascular and metabolic diseases, respiratory morbidity and mortality, mental health disorders, frailty among older adults, and adverse child health outcomes (H. Wang, Gholami, et al., 2024). Together, this broader body of evidence suggests that the observed associations between greenspace morphology and disability in the present study are unlikely to be isolated or outcome specific. Rather, they appear to reflect more general pathways through which the spatial configuration of neighborhood environments shapes physical, mental, and functional health across the life course, ultimately influencing the accumulation or mitigation of disability.

### 4.4 Potential mechanism linking greenspace to disability risk

The relationship between greenspace morphology and disability prevalence may be explained through two primary mechanisms. First, ecological functions likely play an important role. Characteristics of greenspace morphology, including patch size, connectedness, diversity, and shorter distances between greenspace patches, can shape ecological services such as air pollution mitigation and temperature regulation, both of which are relevant to disability risk. Larger and more connected greenspaces tend to reduce particulate matter concentrations (Kong et al., 2014; Łowicki, 2019) and improve air quality (Li et al., 2023), while clustered and aggregated vegetation can lower land surface temperatures more effectively than dispersed green cover (Kowe et al., 2021; Xie et al., 2013; Zhou et al., 2011). These functions may be especially important because exposure to air pollution has been linked to physiological disruption, chronic disease progression, and increased risk of disability, including mobility disability (Lv et al., 2020), cognitive and interpersonal disability (Lin et al., 2017), and accelerated progression of physical disability among older adults (Weuve et al., 2016).

The arrangement of greenspace may affect how people interact with and benefit from nearby environments in everyday life. Prior research suggests that greenspace morphology may influence health through behavioral pathways such as leisure-time physical activity (H. Wang & Tassinary, 2024), and more recent evidence further shows that leisure-time physical activity may mediate the relationship between greenspace morphology and disability prevalence (Xu & Wang, 2025). This mechanism is plausible in the present study because physical activity, routine outdoor exposure, and social interaction are closely related to the development and maintenance of functional health across the life course. For children, diverse and connected greenspaces may create more opportunities for play, exploration, and social engagement. For older adults, well-connected and spatially proximate greenspaces may support easier navigation, walking, and restorative outdoor experiences, including among those using mobility aids. More diverse greenspaces may also accommodate a wider range of preferences, abilities, and everyday uses (de Keijzer et al., 2020; Ekkel & de Vries, 2017). Further research is needed to examine how behavioral processes, including physical activity, social interaction, and daily mobility patterns, may mediate or moderate the relationship between greenspace morphology and disability prevalence across age groups.

### 4.5 Practical implications for health and urban planning

Several practical strategies follow these findings. Enhancing the amount, continuity, and connectivity of neighborhood greenspace may help support a wider range of everyday activities and needs across children, working-age adults, and older adults. Expanding urban open spaces through parks, wooded areas, and other vegetated settings may strengthen opportunities for recreation, restoration, and social interaction. Establishing linked green routes through streetscapes, linear plantings, or small connector spaces may help reduce the isolation of existing green areas and improve continuity at the neighborhood level. In places where vegetated areas are currently scattered or disconnected, modest interventions such as tree planting, vegetated medians, or small linking landscape elements may help reinforce spatial connections among patches while remaining feasible within existing urban fabrics. From a planning perspective, the value of urban greenspace depends not only on whether green areas are present, but also on how they are arranged within the broader neighborhood system and how easily they can be reached and used in everyday life (Endalew Terefe & Hou, 2024; Przewoźna et al., 2024). In addition, given evidence that neighborhoods with higher disability prevalence often have less access to greenspace (Lasky et al., 2023), planning interventions should be especially attentive to underserved communities and should aim to create urban greenspace systems that are not only more abundant, but also more connected, diverse, and accessible across the life course.

### 4.6 Limitations and future research directions

Several limitations should be considered when interpreting the findings. Although the ecological design provides valuable evidence for planning and public health purposes, the cross-sectional nature of the analysis does not permit causal inference and limits our ability to identify underlying mechanisms with certainty. The study also lacks direct measures of individuals’ actual exposure to greenspace, including how frequently people across different age groups visited, used, or interacted with nearby green environments. Future studies using individual-level, longitudinal, or activity-space-based data would help clarify how greenspace morphology shapes actual exposure to nature and influences disability-related outcomes across the life course. Another limitation is that disability was examined as a broad category across age groups. While our broad disability definition provides a stable and policy-relevant measure at the census-tract level, the mechanisms linking greenspace morphology to disability may differ across disability domains, such as mobility, cognitive, sensory, or self-care limitations. We were not able to do this because of the limited sample size when analyzed separately for each disability domain. Future studies should investigate specific disability domains and age-specific pathways when data supports such analysis.

## 5. Conclusions

This study provides national-scale evidence that the spatial morphology of neighborhood greenspace adds value to understanding disability prevalence beyond overall greenspace abundance, underscoring the importance of how green environments are structured rather than merely how much green space is present. We find that multiple greenspace morphological characteristics, including larger patch size, higher patch density, greater connectedness, shorter inter-greenspace distance, greater shape complexity, and higher compositional diversity, were associated with lower disability prevalence across multiple age groups, with more consistent and pervasive associations observed among children and older adults than among working-age adults. These findings suggest that urban planning and landscape design efforts should prioritize neighborhoods with higher disability prevalence and explicitly consider existing greenspace configurations, focusing on reshaping green systems toward more connected, appropriately sized, and functionally supportive morphologies rather than relying on uniform greening strategies.

Finally, we posit that the relationship between greenspace morphology and disability is likely shaped by multiple, interacting mechanisms rather than a single pathway, with environmental processes such as air pollution mitigation and behavioral pathways related to physical activity, alongside impacts on both mental and physical health, jointly contributing to observed population-level patterns of disability.

## Data Availability

All data produced in the present study are available upon reasonable request to the authors

