## Supplementary Materials for "Beyond greenness: Greenspace morphology associates with disability prevalence among children, working-age adults, and older adults—a nationwide study"

**For article:**

Table 1. Lagrange test for modeling disability of older adults

| Model | SEM_AIC | Lag_AIC | SEM_lambda | Lag_rho | SEM_resid_Moran_I | SEM_resid_Moran_p | Lag_resid_Moran_I | Lag_resid_Moran_p | Delta_AIC_Lag_minus_SEM | Preferred_by_AIC |
|---|---|---|---|---|---|---|---|---|---|---|
| PLAND | 470,091.4 | 469,943.7 | 0.246 | 0.240 | -0.008 | 0.999 | -0.006 | 0.995 | -147.660 | Lag |
| PD | 470,032.8 | 469,896.4 | 0.244 | 0.237 | -0.008 | 0.999 | -0.006 | 0.992 | -136.371 | Lag |
| AREA_MN | 470,069.1 | 469,917.8 | 0.246 | 0.240 | -0.008 | 0.999 | -0.006 | 0.995 | -151.348 | Lag |
| COHESION | 470,088.2 | 469,945.7 | 0.247 | 0.240 | -0.008 | 0.999 | -0.006 | 0.995 | -142.470 | Lag |
| SHAPE_AM | 470,031.6 | 469,905.5 | 0.245 | 0.238 | -0.008 | 0.999 | -0.006 | 0.990 | -126.053 | Lag |
| ENN_MN | 470,036.6 | 469,905.9 | 0.244 | 0.238 | -0.008 | 0.999 | -0.006 | 0.990 | -130.632 | Lag |
| SHDI | 470,088.8 | 469,942.2 | 0.246 | 0.240 | -0.008 | 0.999 | -0.006 | 0.995 | -146.522 | Lag |

Note. Delta AIC was calculated as Lag AIC minus SEM AIC; negative values indicate better fit for the spatial lag model, whereas positive values indicate better fit for the spatial error model. Lambda represents the spatial error parameter in SEM, and rho represents the spatial autoregressive parameter in the spatial lag model. Moran’s I tests assess residual spatial autocorrelation. Across all older-adult models, consistently negative ΔAIC values indicate strong support for the spatial lag specification.

Table 2. Lagrange test for modeling disability of working-age adults

| Model | SEM_AIC | Lag_AIC | SEM_lambda | Lag_rho | SEM_resid_Moran_I | SEM_resid_Moran_p | Lag_resid_Moran_I | Lag_resid_Moran_p | Delta_AIC_Lag_minus_SEM | Preferred_by_AIC |
|---|---|---|---|---|---|---|---|---|---|---|
| PLAND | 366,668.1 | 365,620.6 | 0.449 | 0.411 | -0.025 | 1 | -0.019 | 1 | -1,047.499 | Lag |
| PD | 366,669.6 | 365,622.4 | 0.449 | 0.411 | -0.025 | 1 | -0.019 | 1 | -1,047.156 | Lag |
| AREA_MN | 366,665.9 | 365,621.4 | 0.449 | 0.411 | -0.025 | 1 | -0.019 | 1 | -1,044.495 | Lag |
| COHESION | 366,670.0 | 365,606.7 | 0.449 | 0.412 | -0.025 | 1 | -0.019 | 1 | -1,063.344 | Lag |
| SHAPE_AM | 366,572.6 | 365,601.1 | 0.448 | 0.410 | -0.025 | 1 | -0.018 | 1 | -971.486 | Lag |
| ENN_MN | 366,652.3 | 365,597.5 | 0.449 | 0.411 | -0.025 | 1 | -0.019 | 1 | -1,054.880 | Lag |
| SHDI | 366,664.9 | 365,595.8 | 0.449 | 0.411 | -0.025 | 1 | -0.019 | 1 | -1,069.148 | Lag |

Note. Delta AIC was calculated as Lag AIC minus SEM AIC; negative values indicate better fit for the spatial lag model, whereas positive values indicate better fit for the spatial error model. Lambda represents the spatial error parameter in SEM, and rho represents the spatial autoregressive parameter in the spatial lag model. Moran's I tests assess residual spatial autocorrelation. Across all adult models, large negative ΔAIC values indicate strong support for the spatial lag specification.

Table 3. Lagrange test for modeling disability of children

| Model | SEM_AIC | Lag_AIC | SEM_lambda | Lag_rho | SEM_resid_Moran_I | SEM_resid_Moran_p | Lag_resid_Moran_I | Lag_resid_Moran_p | Delta_AIC_Lag_minus_SEM | Preferred_by_AIC |
|---|---|---|---|---|---|---|---|---|---|---|
| PLAND | 65,301.83 | 65,284.75 | 0.051 | 0.062 | 0.007 | 0.102 | -0.001 | 0.569 | -17.078 | Lag |
| PD | 65,272.66 | 65,257.68 | 0.050 | 0.060 | 0.007 | 0.121 | -0.001 | 0.565 | -14.976 | Lag |
| AREA_MN | 65,276.17 | 65,260.94 | 0.051 | 0.061 | 0.007 | 0.120 | -0.001 | 0.566 | -15.229 | Lag |
| COHESION | 65,274.21 | 65,259.05 | 0.050 | 0.060 | 0.007 | 0.119 | -0.001 | 0.566 | -15.165 | Lag |
| SHAPE_AM | 65,303.54 | 65,286.69 | 0.051 | 0.062 | 0.007 | 0.105 | -0.001 | 0.569 | -16.846 | Lag |
| ENN_MN | 65,261.73 | 65,246.61 | 0.050 | 0.060 | 0.007 | 0.117 | -0.001 | 0.563 | -15.119 | Lag |
| SHDI | 65,283.04 | 65,265.98 | 0.051 | 0.061 | 0.007 | 0.100 | -0.001 | 0.568 | -17.053 | Lag |

Note. Delta AIC was calculated as Lag AIC minus SEM AIC; negative values indicate better fit for the spatial lag model, whereas positive values indicate better fit for the spatial error model. Lambda represents the spatial error parameter in SEM, and rho represents the spatial autoregressive parameter in the spatial lag model. Moran's I tests assess residual spatial autocorrelation. Across all adult models, large negative ΔAIC values indicate strong support for the spatial lag specification.

**Example of greenspace classification and morphology metrics in the Atlanta metropolitan area**

To provide a visual illustration of the greenspace classification and morphology metrics used in the national-scale analyses, Appendix B presents an example from the Atlanta metropolitan area. Atlanta was selected because it contains substantial spatial variation in urban development intensity, greenspace distribution, fragmentation, and connectivity patterns, making it well suited for demonstrating the morphology framework applied throughout this study. The appendix first illustrates the original NLCD 2023 land-cover data and the binary greenspace classification used in the analyses, followed by examples of the tract-level morphology metrics calculated for the study area, including greenspace percentage, patch density, mean patch area, cohesion, shape complexity, inter-greenspace distance, and landscape diversity. These figures are intended to visually demonstrate how different dimensions of greenspace morphology vary across an urban metropolitan landscape.

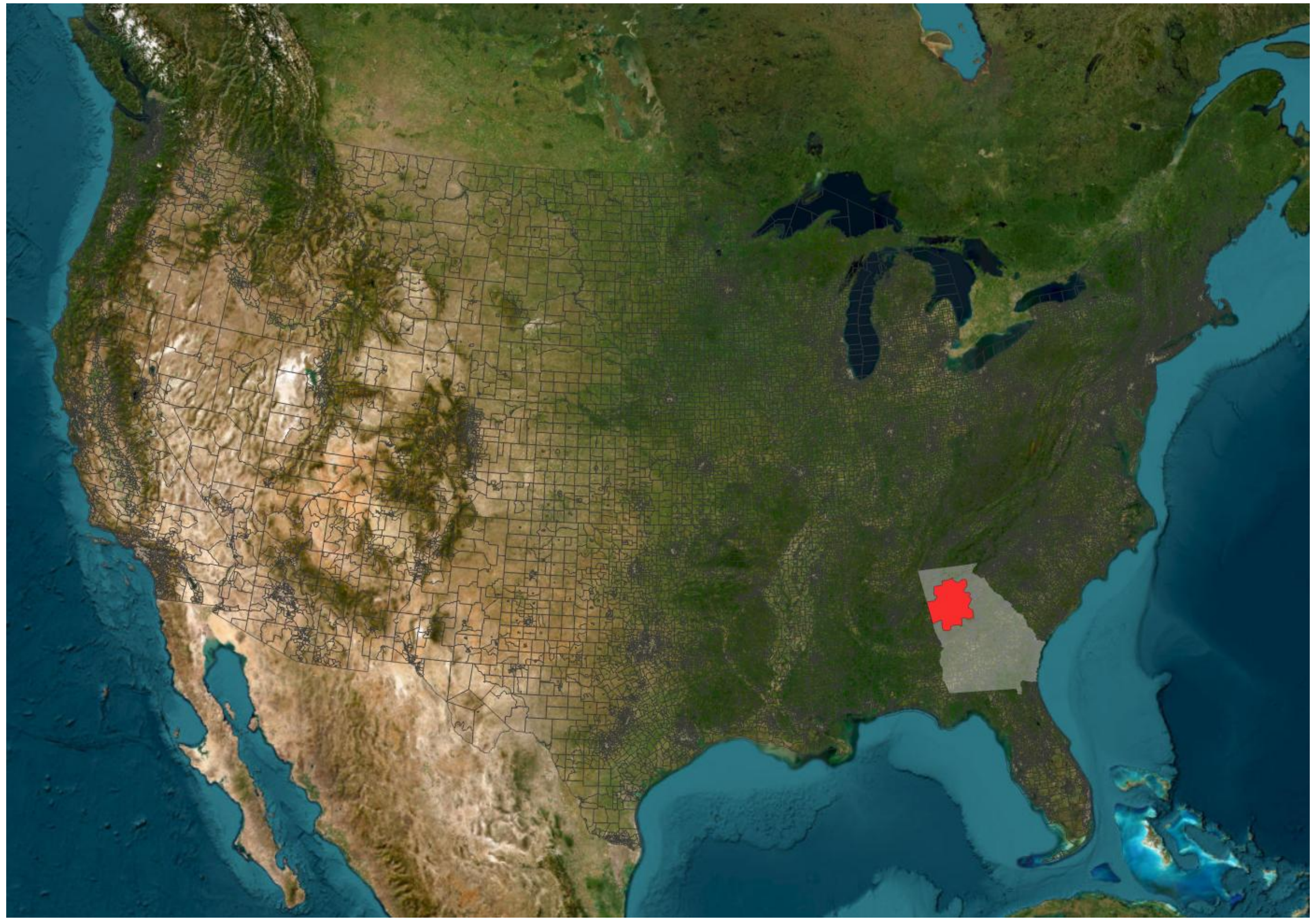

*Location of the Atlanta metropolitan area within the contiguous United States. The Atlanta metropolitan area was selected as an illustrative example to demonstrate the greenspace classification and morphology metrics used in the study.*

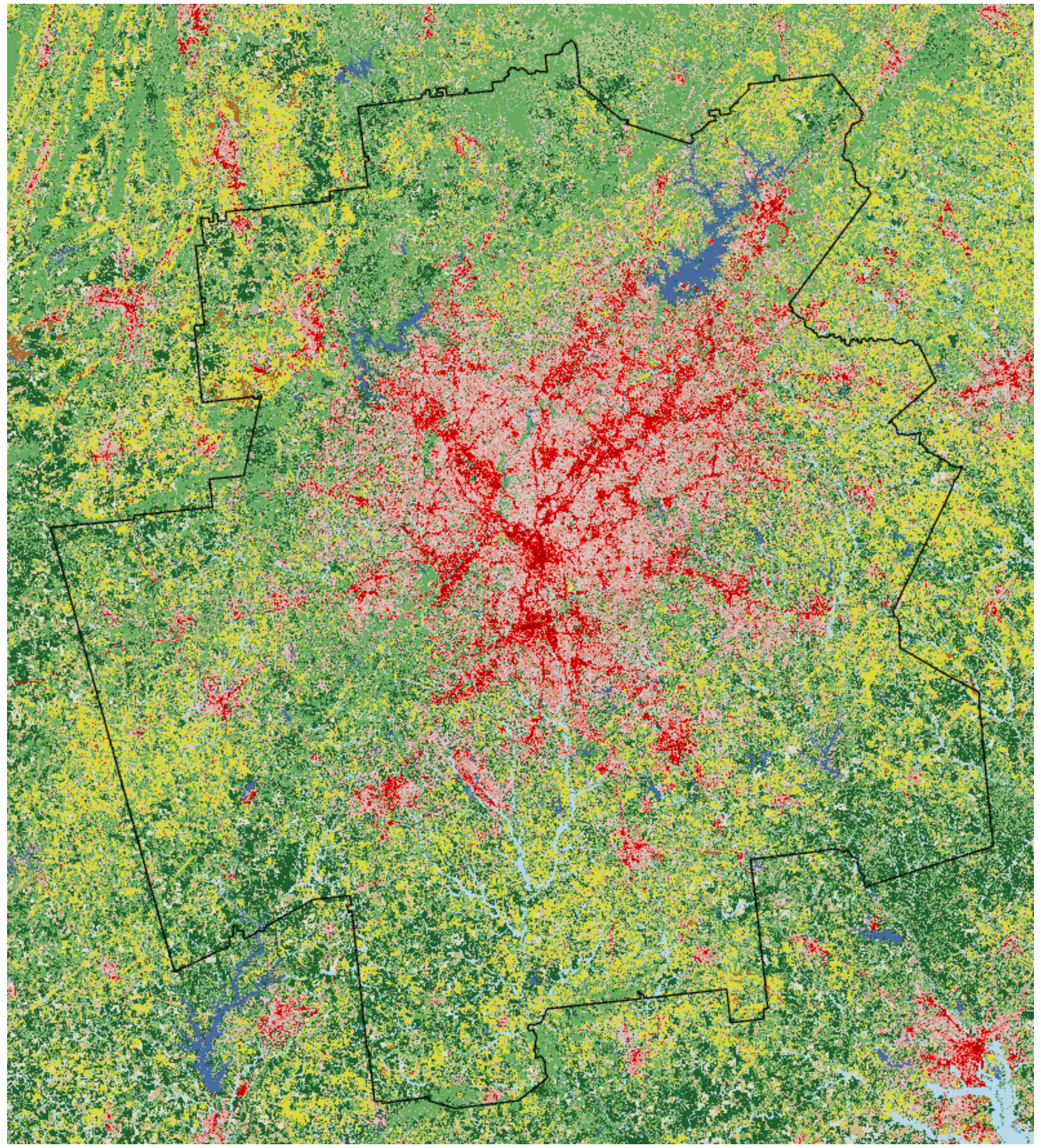

(a) Original NLCD 2023 land-cover classification

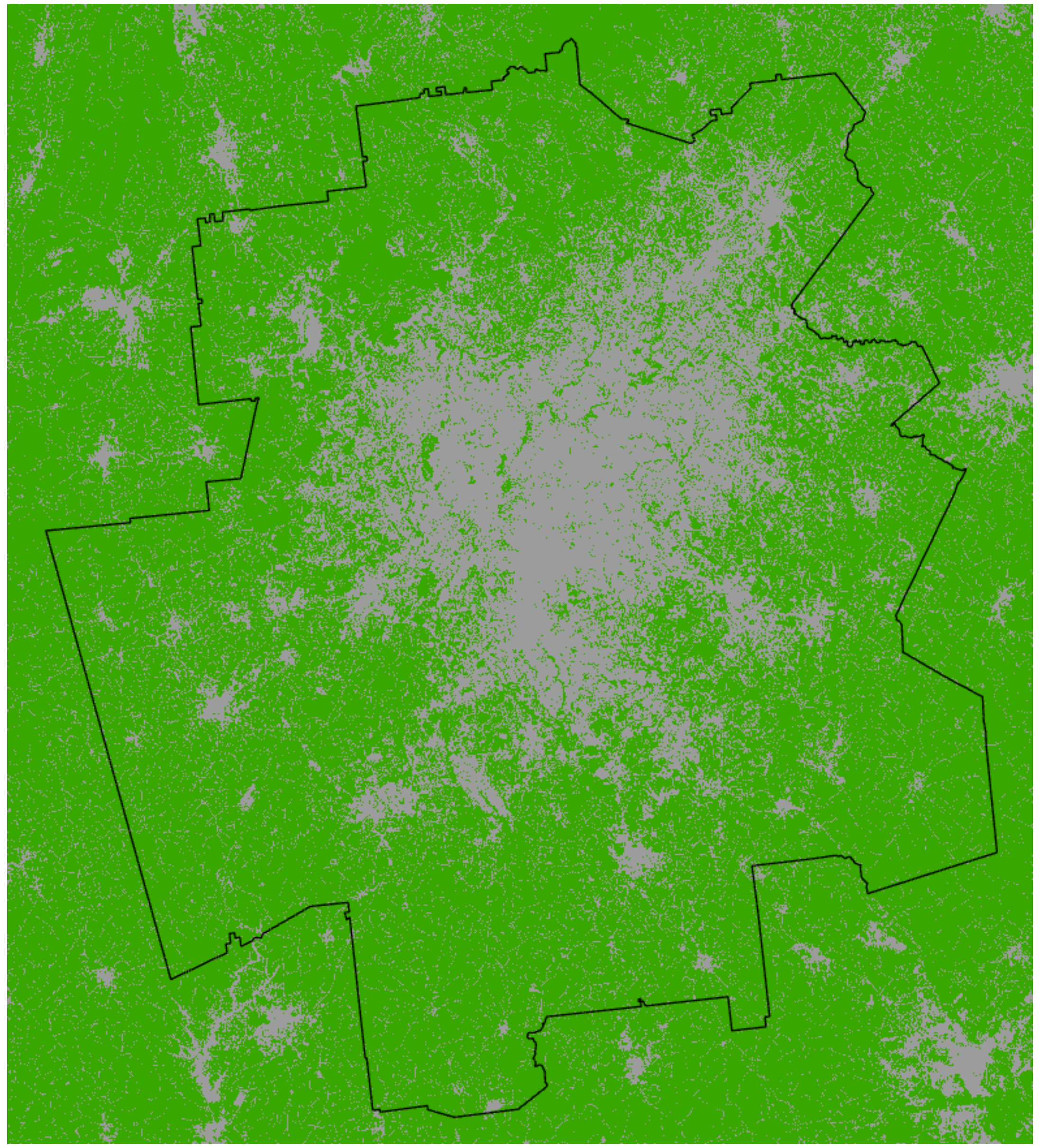

(b) Binary greenspace classification map
Green = greenspace
Gray = non-greenspace

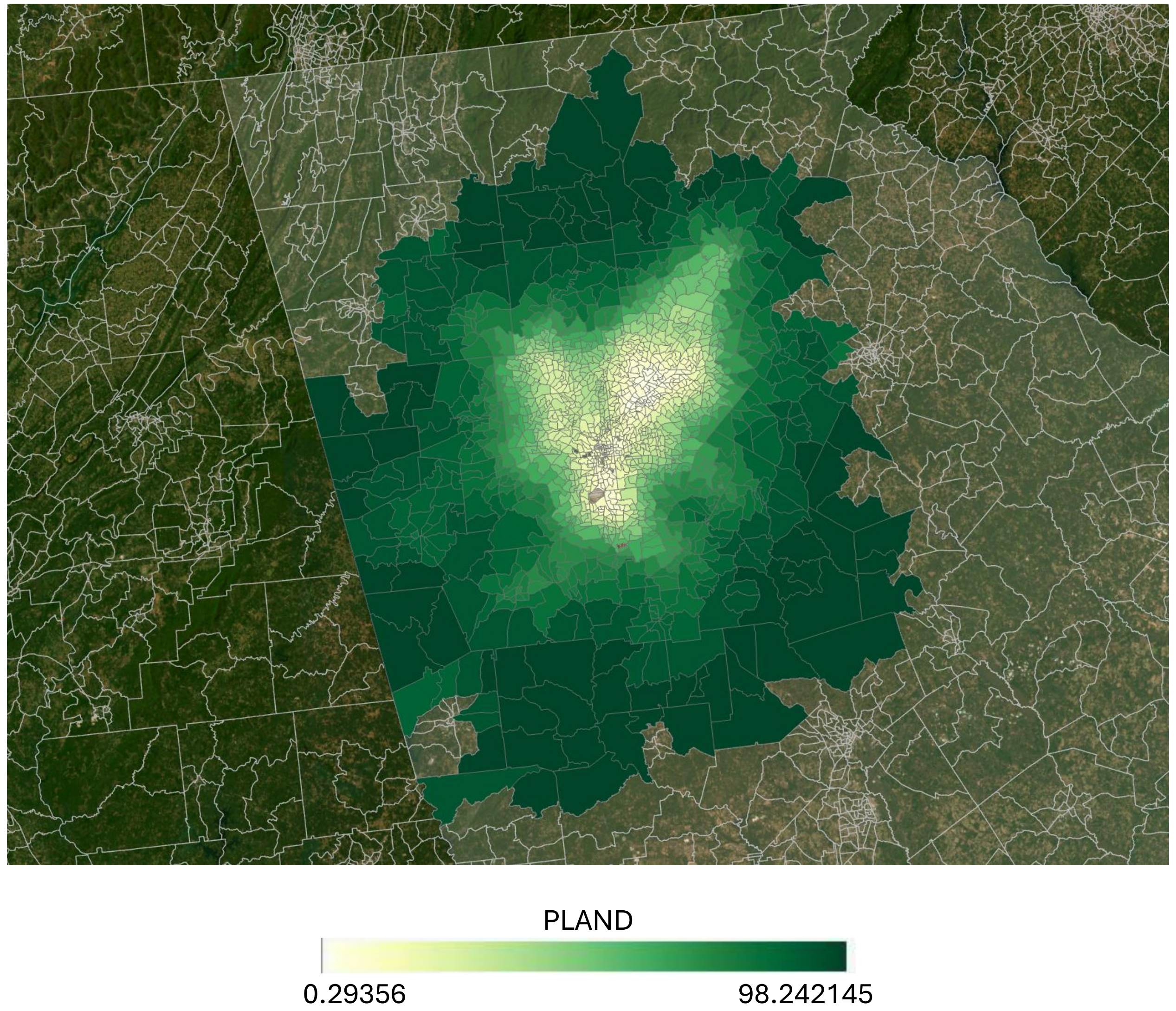
PLAND
0.29356
98.242145

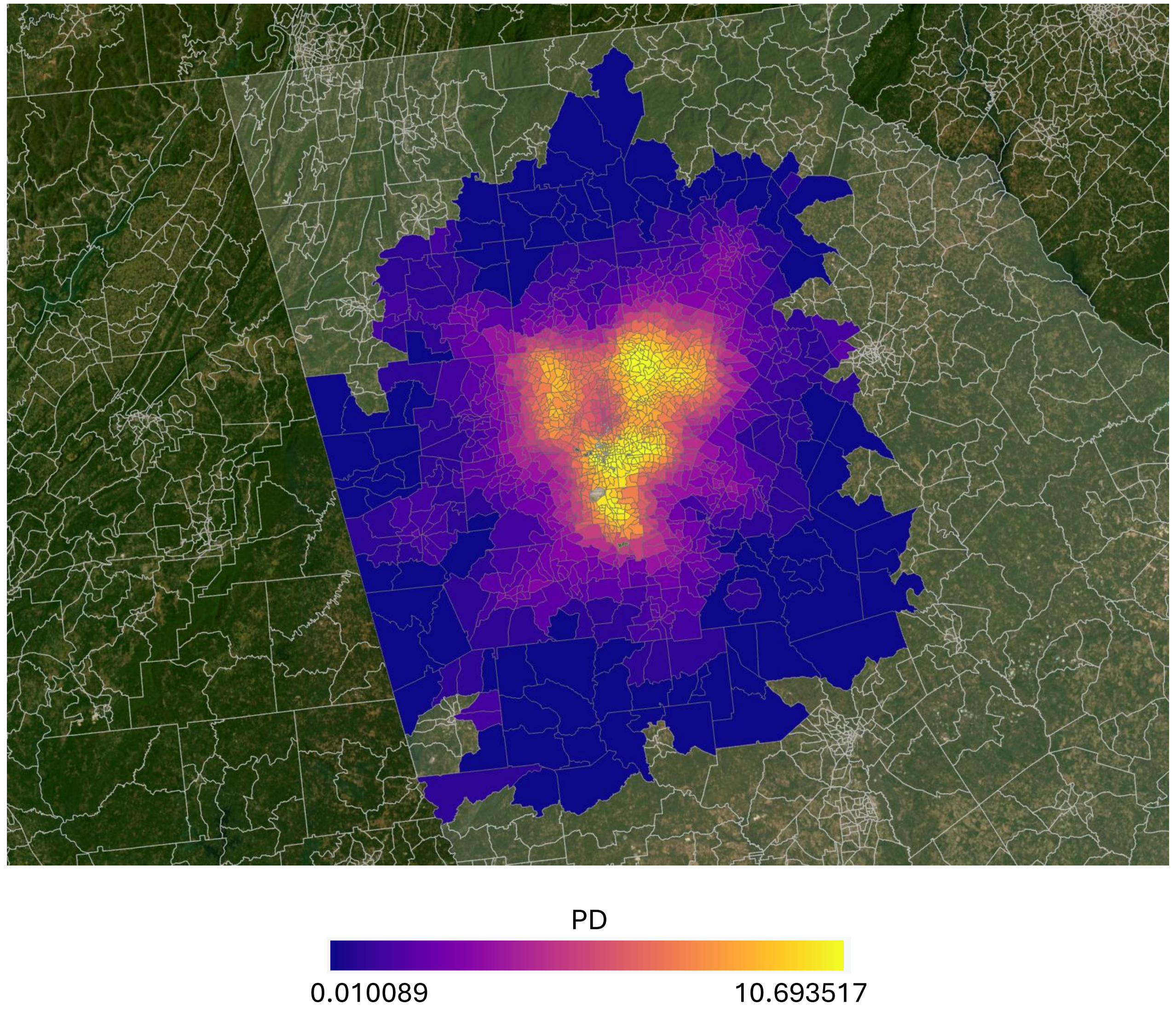
PD
0.010089
10.693517

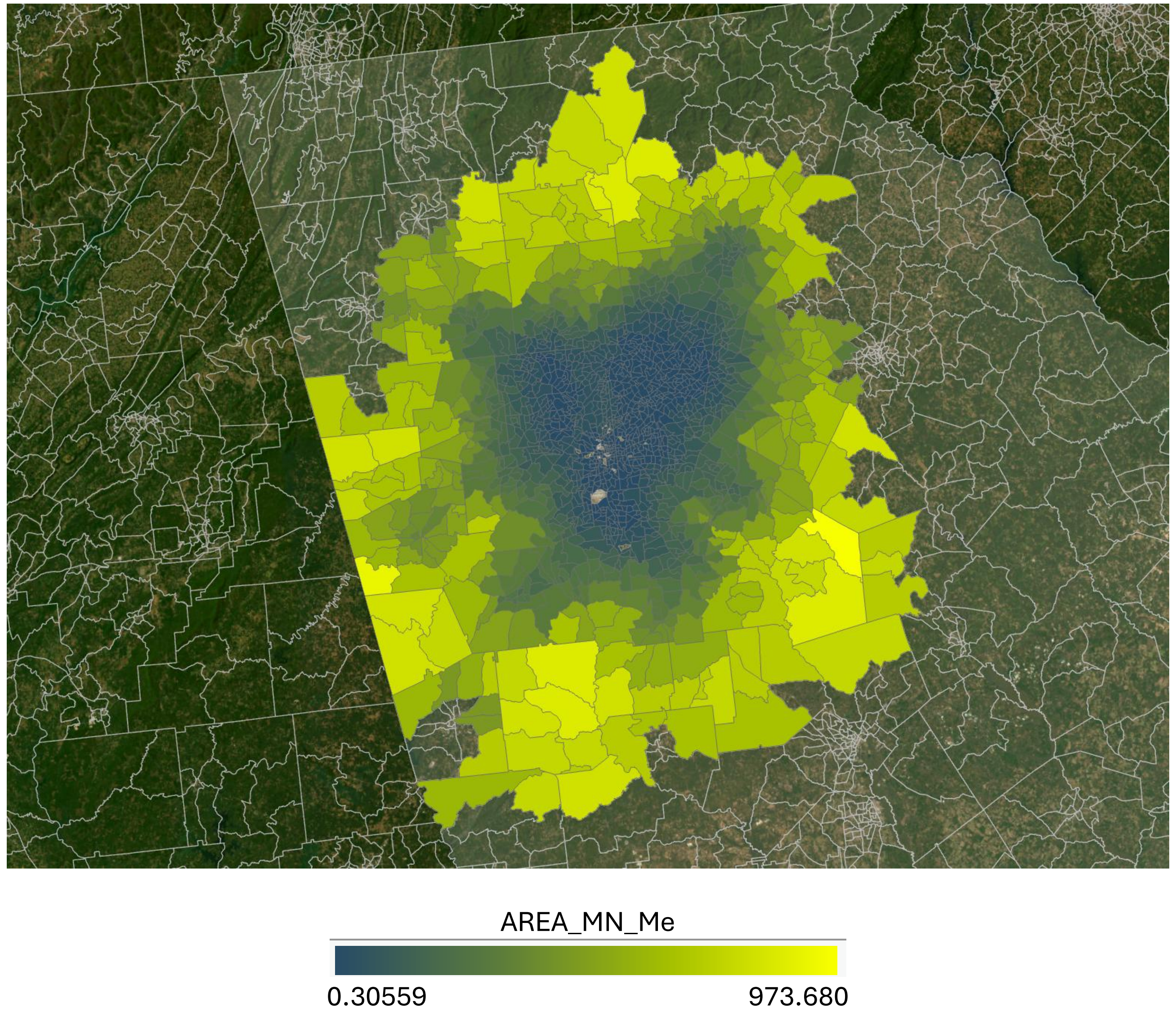
AREA_MN_Me
0.30559
973.680

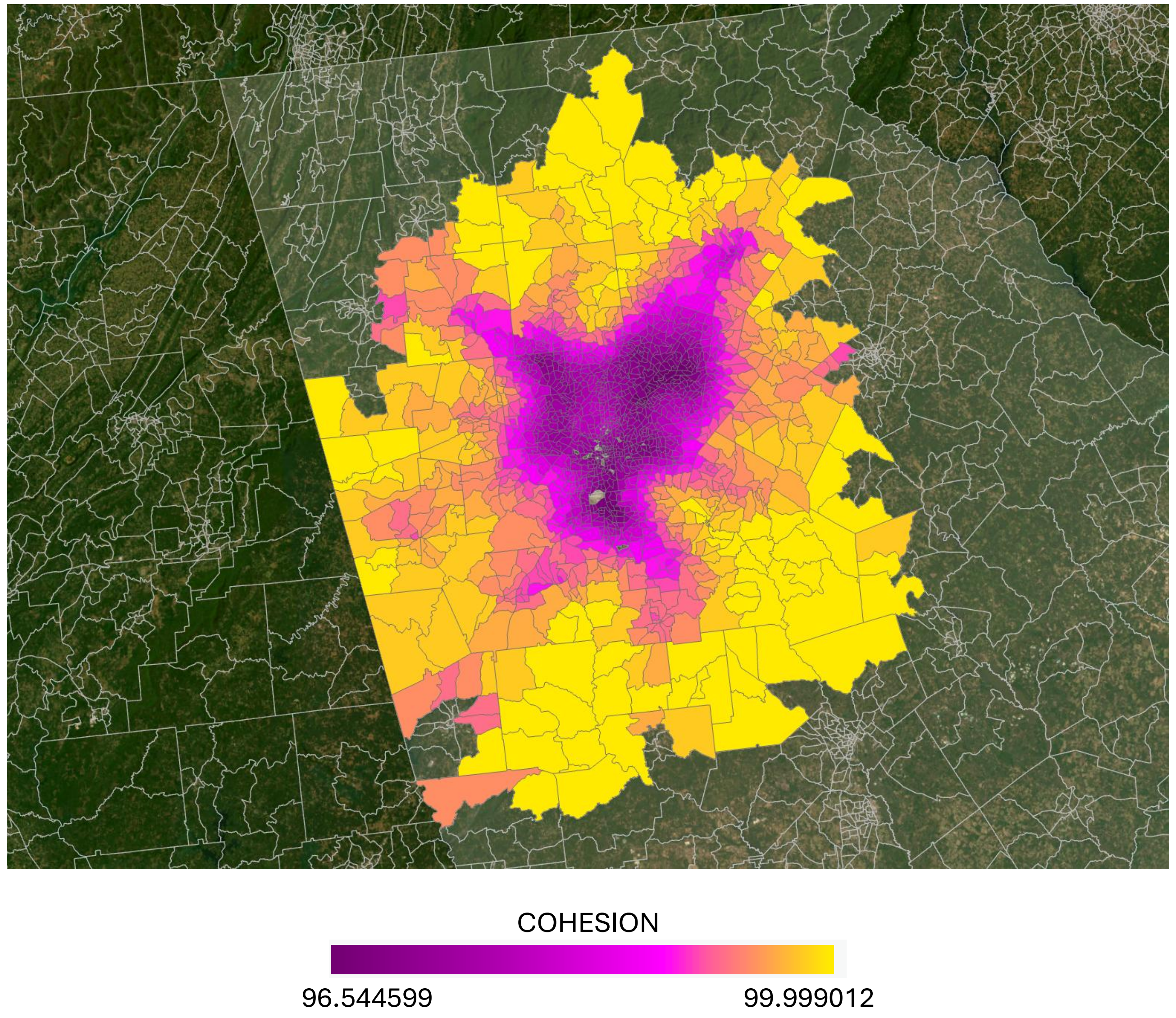
COHESION
96.544599
99.999012

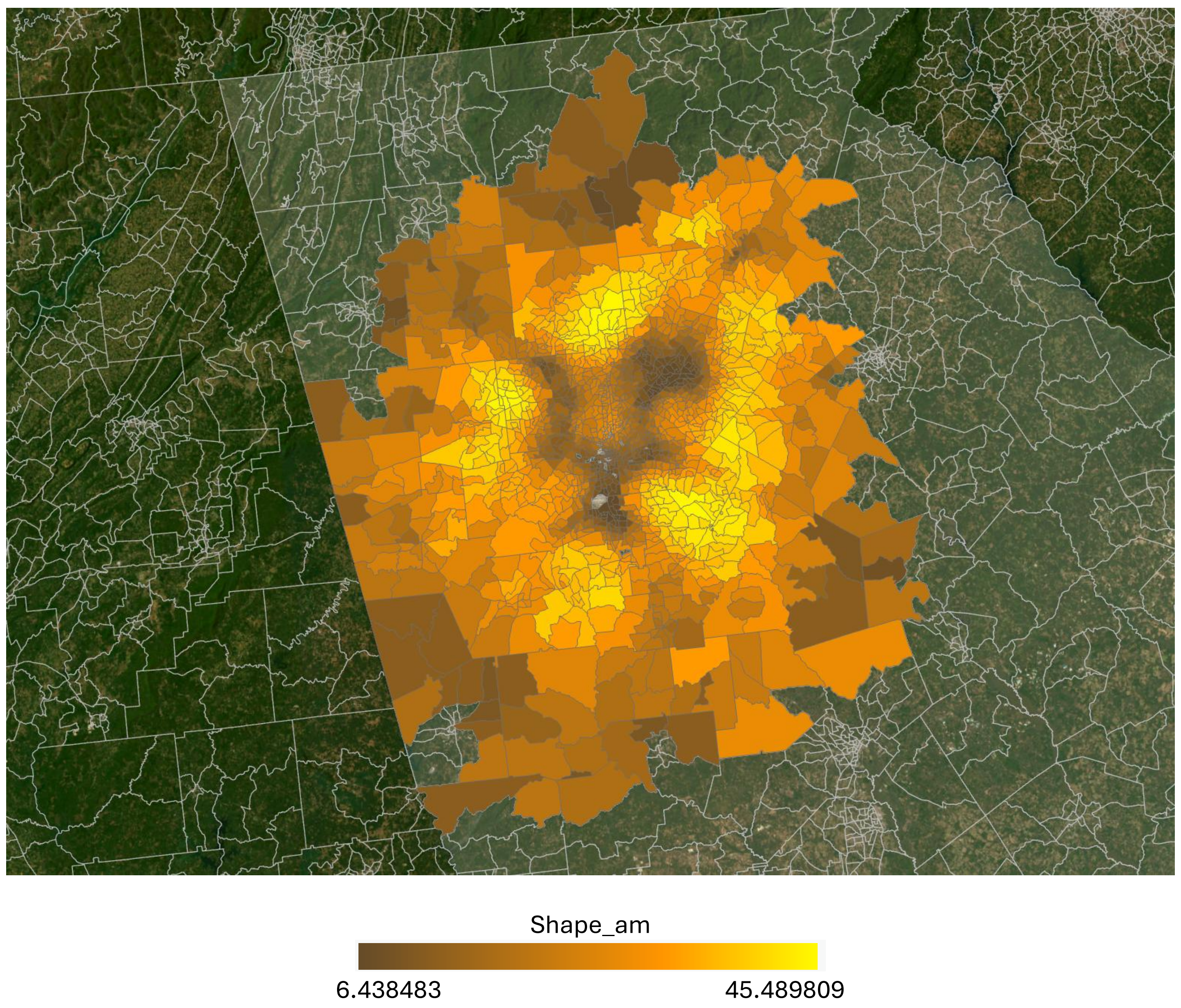
Shape_am
6.438483
45.489809

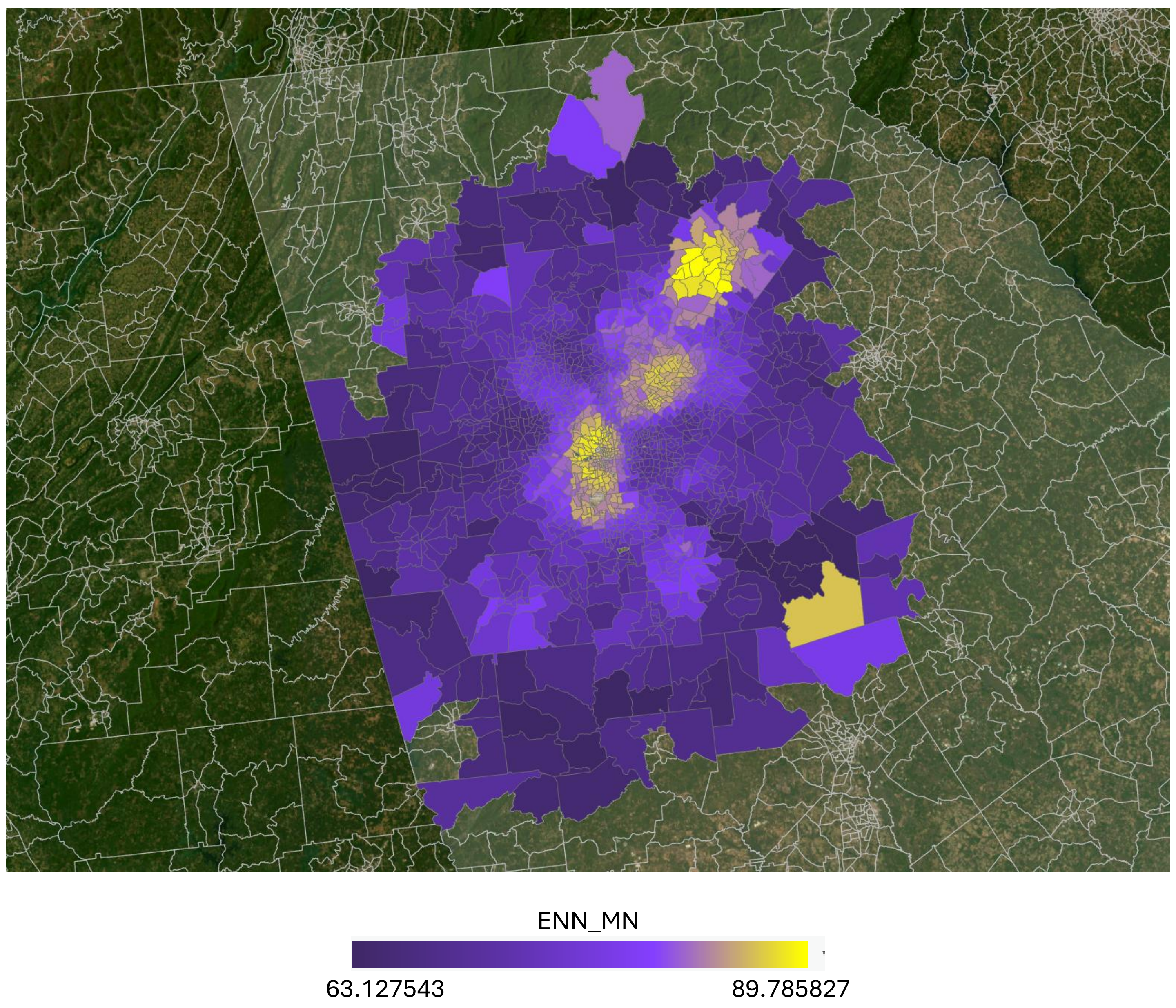
ENN_MN
63.127543
89.785827

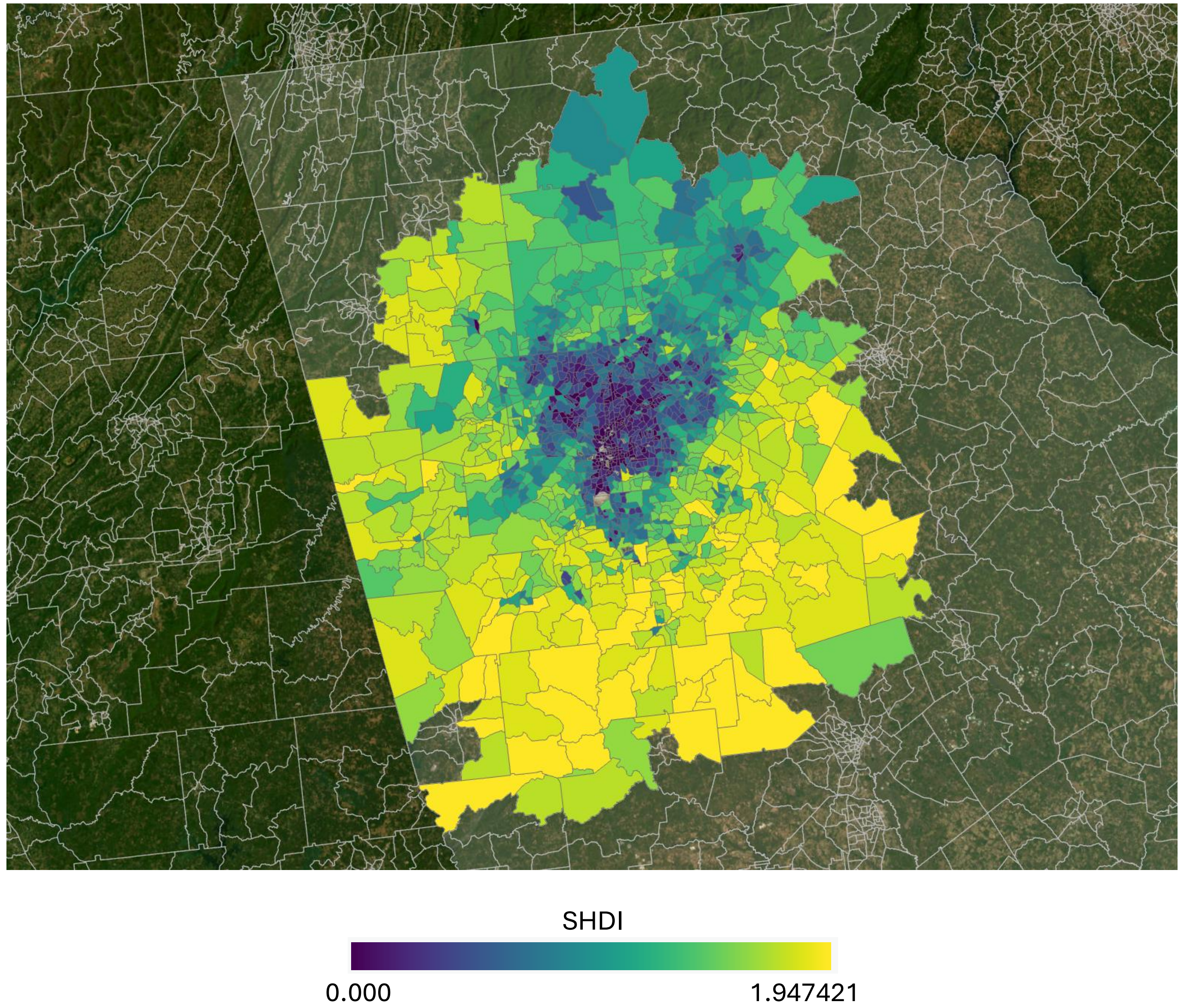
SHDI
0.000
1.947421
